# CCL17-CCR4 Signalling Attenuates Pressure Overload-Induced Cardiac Remodelling Through IGFBP6 Suppression in Cardiac Fibroblasts

**DOI:** 10.64898/2026.11.11.26367804

**Authors:** Wendi Yan, Pingjing Zheng, Zhipeng Lian, Gaocheng Gu, Xin Wang, Xin Chen, Chun-Ka Wong, Yang Zhang, Wei Guo, Yuxiang Dai

## Abstract

**Background and Aims:** Chemokines are key regulators of immune-cell trafficking, inflammation, and tissue repair in cardiovascular disease. C-C motif chemokine ligand 17 (CCL17) has been linked to adverse remodelling after myocardial infarction and neurohumoral stress, but its function during sustained mechanical pressure overload is unknown. We investigated whether CCL17 regulates ventricular remodelling through cardiac fibroblast signalling.

**Methods:** Serum proteomic screening and myocardial and serum validation were performed in patients with aortic stenosis and healthy controls. CCL17 function was evaluated using global *Ccl17* deletion and adeno-associated virus-mediated CCL17 augmentation in mice subjected to transverse aortic constriction. Cardiac transcriptomics and complementary pharmacological, small interfering RNA, and promoter-reporter experiments in cardiac fibroblasts were used to define downstream signalling. Separately, fibroblast-directed *Igfbp6* silencing was used to test the functional contribution of IGFBP6 in vivo.

**Results:** Serum CCL17 concentrations and myocardial CCL17 abundance were increased in patients with aortic stenosis, and cardiac and circulating CCL17 increased rapidly after transverse aortic constriction. *Ccl17* deletion aggravated pressure overload-induced systolic dysfunction, hypertrophy, and fibrosis, whereas CCL17 augmentation conferred reciprocal protection. Transcriptomic analyses identified insulin-like growth factor-binding protein 6 (IGFBP6) as a CCL17-responsive candidate. In cardiac fibroblasts, CCL17 reduced secreted and cellular IGFBP6 and α-smooth muscle actin through C-C chemokine receptor 4 (CCR4)-dependent signalling associated with suppression of early growth response 1 (EGR1). Genetic perturbation and promoter-reporter analyses further supported CCR4–EGR1-dependent regulation of *Igfbp6*. Finally, fibroblast-directed *Igfbp6* silencing improved systolic function and reduced hypertrophy and fibrosis in *Ccl17*-deficient mice after transverse aortic constriction.

**Conclusions:** Our findings identify CCL17 as a previously unrecognized cardioprotective factor and uncover a novel immune cell–fibroblast communication axis, whereby CCL17 restrains pressure overload–induced maladaptive cardiac remodelling through CCR4–EGR1–mediated suppression of IGFBP6 in cardiac fibroblasts.

**Graphical abstract:** 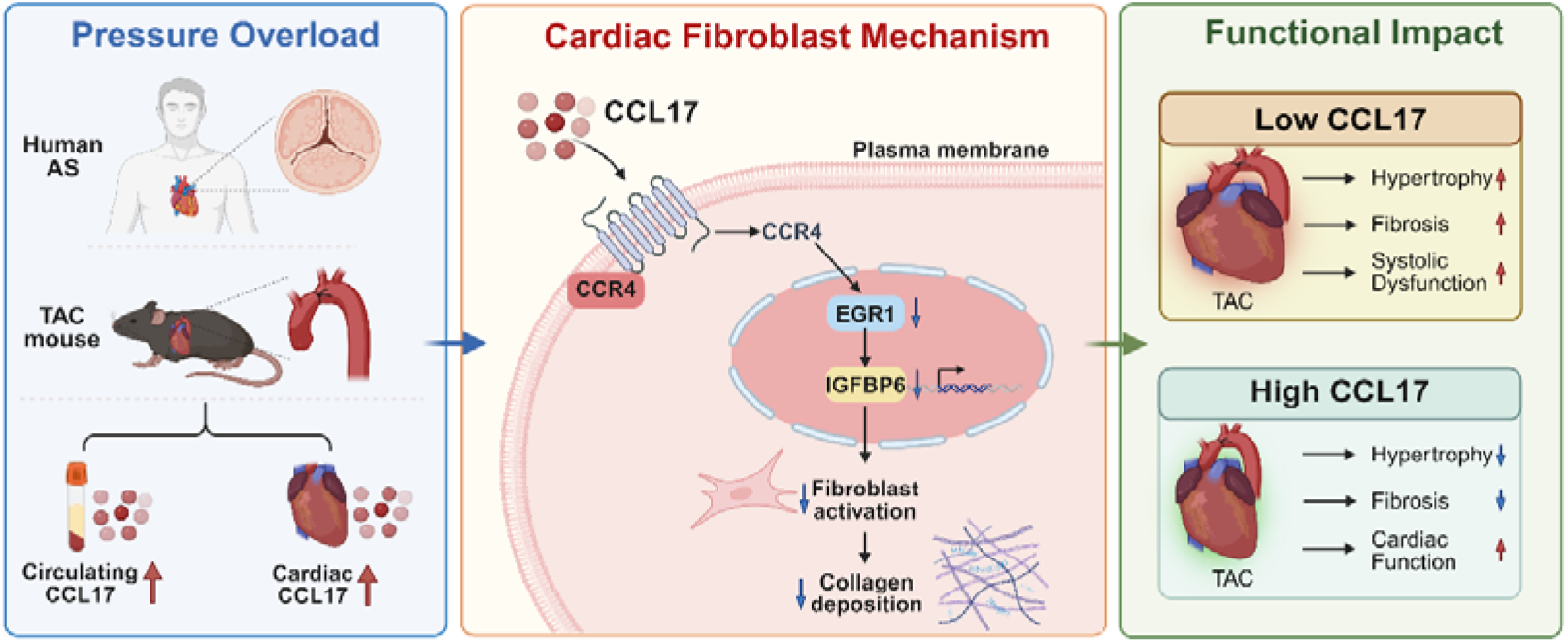

Pressure overload was associated with increased circulating and myocardial CCL17 in patients with aortic stenosis and mice subjected to transverse aortic constriction. In cardiac fibroblasts, CCL17 signals through CCR4 to suppress EGR1-dependent Igfbp6 transcription, thereby restraining fibroblast activation and extracellular matrix deposition. Accordingly, Ccl17 deficiency aggravated pressure overload-induced cardiac hypertrophy, fibrosis, and systolic dysfunction, whereas AAV-mediated CCL17 augmentation conferred reciprocal protection. Fibroblast-directed Igfbp6 silencing partially rescued the adverse remodelling phenotype in Ccl17-deficient mice. These findings identify CCL17 as an endogenous protective signal and establish the CCR4–EGR1–IGFBP6 pathway as a functionally relevant regulator of fibroblast activation during pressure overload.

## Introduction

Pressure overload elicits both adaptive and maladaptive responses in the left ventricle. With sustained stress, this balance may shift towards hypertrophy, fibrosis, and cardiac dysfunction. Aortic stenosis (AS) provides a clinically relevant setting in which to investigate these responses, as progressive outflow obstruction is accompanied by myocardial remodelling ^[1–3]^. Recent human multi-omic analyses have linked circulating proteins to myocardial transcriptional programmes and remodelling phenotypes in AS ^[2]^. Although these associations can nominate biologically relevant candidates, they cannot determine whether an individual signal drives myocardial injury or represents a compensatory response that limits it. Defining the functional roles of these signals is therefore essential for identifying endogenous protective mechanisms that support ventricular adaptation to pressure overload.

Cardiac fibroblasts are key effectors of myocardial remodelling and potential recipients of stress-responsive signals. Lineage-tracing studies have identified resident fibroblasts as a major source of activated myofibroblasts in the injured heart ^[4]^. Their activation is governed by distinct transcriptional programmes that are amenable to pharmacological modulation^[5]^ and is further shaped by signals from immune cells. In failing hearts, macrophage-derived interleukin-1β promotes disease-associated fibroblast states, whereas disruption of this signalling axis attenuates fibrosis in experimental models^[6]^. Together, these findings highlight immune-fibroblast communication as an important regulator of cardiac remodelling.^[7]^ However, which immune-derived signals, if any, restrain excessive fibroblast activation during sustained pressure overload remains unclear.

Chemokines coordinate immune-cell trafficking and inflammatory responses during cardiac injury and can influence subsequent cardiac remodelling^[8–10]^. Among these, C-C motif chemokine ligand 17 (CCL17) has been implicated in several models of cardiac injury. Feng et al. showed that CCL17 impaired regulatory T-cell recruitment and exacerbated cardiac remodelling after myocardial infarction or combined angiotensin II and phenylephrine infusion^[8]^. Consistent with these findings, our previous work demonstrated that CCL17 promotes maladaptive remodelling in models of ageing-associated and angiotensin II-induced cardiac hypertrophy^[9]^. These studies have largely attributed the effects of CCL17 to immune-cell regulation^[11, 12]^, including CCR4-dependent signalling^[13]^. Whether CCL17 has similar effects during pressure overload or acts directly on cardiac fibroblasts remains unknown.

Immune-fibroblast communication may extend beyond inflammatory signalling to programmes governing tissue growth and repair. The insulin-like growth factor (IGF) system represents one such regulatory network, in which IGF-binding proteins (IGFBPs) modulate IGF bioavailability and can also exert IGF-independent effects^[15]^. Recent work identified fibroblast-derived IGFBP6 as a promoter of myofibroblast activation and adverse cardiac remodelling after myocardial infarction or isoproterenol exposure, involving an interaction with early growth response 1 (EGR1) and downstream MFAP4 transcription^[16]^. Whether immune-derived signals regulate IGFBP6 expression and thereby influence fibroblast activation during sustained pressure overload remains unknown.

Here, we investigated whether CCL17, a chemokine previously linked to adverse cardiac remodelling, might exert a distinct function during sustained mechanical pressure overload. Integrating human AS proteomics and myocardial analyses with complementary loss-and gain-of-function studies in TAC mice, we identified CCL17 as an endogenous protective factor that limits pathological cardiac hypertrophy, fibrosis and ventricular dysfunction. Mechanistic studies in cardiac fibroblasts, together with fibroblast-directed *Igfbp6* silencing, support a CCL17-CCR4-EGR1-IGFBP6 signalling pathway that restrains fibroblast activation and adverse remodelling. These findings reveal an unexpected cardioprotective role for CCL17 and highlight chemokine-fibroblast communication as a link between immune-associated signals and growth-factor-related regulatory programmes in the pressure-overloaded heart.

## Methods

### Study design and human samples

This study integrated human myocardial and serum samples, murine models of pressure overload, and primary cardiac fibroblast experiments to determine the expression, function, and downstream mechanisms of CCL17 during pressure-overload remodelling. All participants provided written informed consent. The study conformed to the Declaration of Helsinki and was approved by the ethics committees of Zhongshan Hospital, Fudan University (B2021-418R), and the First Affiliated Hospital of Guangxi Medical University (2017KY-E-105). Myocardial tissue was obtained from patients with end-stage severe aortic stenosis (AS) undergoing heart transplantation (n=8) and from non-AS surgical controls undergoing valve surgery for isolated mitral or aortic regurgitation (n=8). Samples rinsed in ice-cold saline, snap-frozen in liquid nitrogen, and stored at −80 °C. Severe AS was defined echocardiographically by an aortic valve area ≤1.0 cm², a mean transvalvular gradient ≥40 mmHg, or a peak aortic jet velocity ≥4.0 m/s, together with evidence of left ventricular hypertrophy and dysfunction.^[17]^Exclusion criteria comprised severe hepatic or renal dysfunction, malignant disease, acute or chronic infection, genetically confirmed primary hypertrophic cardiomyopathy, and incomplete clinical data.

The serum discovery cohort initially included 35 patients with AS and 35 healthy controls. One sample in each group was excluded because of platform quality-control warnings, leaving 34 participants per group for analysis. An independent enzyme-linked immunosorbent assay (ELISA) validation cohort comprised 80 patients with AS and 80 healthy controls. Fasting venous blood was allowed to clot for 30 min at room temperature and centrifuged at 1,000 × g for 20 min. Serum was aliquoted, stored at −20 °C for short-term use and at −80 °C for long-term storage, and protected from repeated freeze-thaw cycles. Healthy controls were recruited from a health-screening centre and had no known structural heart disease, coronary artery disease, heart failure, malignant disease, severe hepatic or renal dysfunction, or active infection.

### Olink proteomic profiling

Serum proteins were measured using the Olink Target 96 Immuno-Oncology panel (Olink Proteomics AB, Uppsala, Sweden) according to the manufacturer’s instructions. This proximity extension assay measures 92 proteins from 1 μL of sample: pairs of oligonucleotide-labelled antibodies generate an amplifiable DNA reporter sequence after binding their target protein, and the reporter is quantified on a Signature Q100 microfluidic real-time PCR instrument (LC-Bio Technology, Hangzhou, China).^[18]^ Results were reported as normalized protein expression (NPX), an arbitrary relative unit on a log scale.Internal incubation, extension, and detection controls and inter-plate controls were used for quality control and normalization. A run passed quality control when the within-run standard deviations of the relevant internal controls were <0.2. Samples deviating by >±0.3 NPX from the plate median for incubation control 2 or the detection control received a quality-control warning and were excluded. Quality-controlled NPX values were analysed using a two-sided Mann–Whitney U test. Exploratory differentially abundant proteins were defined by a nominal P<0.05 and |log fold change|>0.5. Heatmaps were row-standardized by Z score, and receiver-operating characteristic analysis was used to calculate the area under the curve and 95% confidence interval for CCL17.^[19]^

### Animals

Eight-week-old male mice on a C57BL/6J background were used. Wild-type mice were obtained from GemPharmatech (Jiangsu, China). Global Ccl17-knockout mice, provided by Professor Yicong Ye, were generated by Cyagen Biosciences using CRISPR/Cas9-mediated targeting of exons 1–3 of *Ccl17*. Mice were housed under specific-pathogen-free conditions (22±2 °C, 50±10% humidity, 12-h light/dark cycle) with ad libitum access to chow and water. Animal procedures were approved by the Animal Ethics Committee of Zhongshan Hospital, Fudan University.^[20]^

### Genotyping of *Ccl17*-knockout mice

Genomic DNA was extracted from tail biopsies after overnight lysis at 55 °C and analysed by PCR. Cycling conditions were 94 °C for 3 min; 35 cycles of 98 °C for 15 s, 60 °C for 15 s, and 68 °C for 1 min; followed by 68 °C for 15 min. Amplicons were resolved on a 1% agarose gel. Wild-type, homozygous knockout, and heterozygous mice yielded 656-bp, 408-bp, and both bands, respectively. Primer sequences are provided in Supplementary Table 1

### Transverse aortic constriction

Pressure overload was induced by transverse aortic constriction (TAC).^[21]^ Mice were anaesthetized with 1.5% isoflurane, intubated, mechanically ventilated, and subjected to thoracotomy to expose the transverse aorta. A 5-0 absorbable suture was tied around the aorta and a 29-gauge spacer, after which the spacer was removed. Sham-operated mice underwent the same procedure without ligation. Echocardiography and tissue collection were performed 6 weeks later.

### AAV-mediated gene modulation

Recombinant AAV9 vectors were generated by Weizhen Biosciences (Shandong, China). AAV9-F4/80-Ccl17-3×FLAG was used for CCL17 augmentation, with AAV9-F4/80-GFP as control. AAV9-COL1A2-GFP-miR30shIgfbp6 was used for fibroblast-enriched Igfbp6 silencing, with a backbone-matched non-targeting vector as control. Viruses were delivered by tail-vein injection at 1×10¹² vector genomes per mouse 1 week before TAC or sham surgery.

### *Ccl17*-tdTomato reporter mice

*Ccl17*-tdTomato reporter mice on a C57BL/6J background were generated by Shanghai Model Organisms Center, Inc. (CM-KI-242469). The targeted allele contains a tdTomato reporter cassette and an FRT-flanked PGK-Neo selection cassette inserted into the exon 2 region of *Ccl17*. Genotyping was performed by PCR using primers P1 (5′-TCGGAACATTCCATGGGTCC-3′), P2 (5′-GCCTCAGGCTGTGTGTTAGT-3′), and P4 (5′-GTGCCATGCCCCAGGAAC-3′). Wild-type, homozygous, and heterozygous mice yielded a 498-bp product, a 765-bp product, and both products, respectively. Homozygous reporter mice underwent sham surgery or TAC, and hearts were collected before surgery and on postoperative days 1, 3, and 7. Immunofluorescence staining for RFP and α-SMA was used to assess *Ccl17* reporter activity and its spatial distribution within regions of cardiac remodelling.

### Echocardiography

Echocardiography was performed using a Vevo 2100 system (VisualSonics). Anaesthesia was induced with 2% isoflurane and maintained at 1.5%; mice were placed on a heated platform and heart rate was maintained at 450–600 beats/min. Parasternal long-axis, short-axis, and M-mode images at the papillary-muscle level were acquired. Measurements from 3–4 stable cardiac cycles were averaged to calculate left ventricular ejection fraction, fractional shortening, and left ventricular mass.

### Histology and morphometry

Paraffin-embedded cardiac sections were stained with a Masson trichrome kit (G1340, Solarbio) according to the manufacturer’s instructions to assess interstitial and perivascular fibrosis. Wheat germ agglutinin (WGA; I3300, Solarbio; 5–10 μg/mL) was applied for 60 min at room temperature in the dark to delineate cardiomyocyte membranes, followed by DAPI nuclear counterstaining. Images were acquired using identical settings, and left ventricular collagen volume fraction and cardiomyocyte cross-sectional area were quantified using ImageJ.^[22]^

### RNA extraction and RT-qPCR

Total RNA was extracted from tissues or cells using an RNA isolation kit (RN001, Yishan), and cDNA was synthesized using PrimeScript RT Master Mix (RR036A, Takara). Quantitative PCR was performed using Hieff qPCR SYBR Green Master Mix (11203ES03, Yeasen) on a QuantStudio 5 Real-Time PCR System (Applied Biosystems) under the following conditions: 95 °C for 5 min; 40 cycles of 95 °C for 10 s and 60 °C for 30 s; followed by melt-curve analysis. Relative expression was normalized to *Gapdh* and calculated using the 2⁻ΔΔCt method.^[23, 24]^

### Immunoblotting

Tissues or cells were lysed in RIPA buffer containing protease and phosphatase inhibitors. Clarified lysates were quantified by bicinchoninic acid assay, and equal amounts of protein were separated by SDS-PAGE and transferred to polyvinylidene difluoride membranes. Membranes were blocked for 20 min, incubated with primary antibodies overnight at 4 °C, and then with horseradish-peroxidase-conjugated secondary antibodies for 1 h at room temperature. Chemiluminescent signals were detected using a Tanon imaging system and quantified with ImageJ.^[22]^ GAPDH or α-tubulin served as loading controls. Antibody source, catalogue number are provided in Supplementary Table2

### ELISA

Human and mouse serum CCL17 concentrations were measured using human CCL17/TARC (DDN00, R&D Systems) and mouse CCL17/TARC (MCC170, R&D Systems) ELISA kits, respectively. IGFBP6 concentrations in NRCF-conditioned medium were measured using a rat IGFBP-6 ELISA kit (NBP2-80237, Novus Biologicals). Assays were performed according to the manufacturers’ instructions, with sample dilution factors established in pilot experiments. Absorbance was measured at 450 nm, and concentrations were calculated using a four-parameter logistic standard curve and corrected for sample dilution.

### Cardiac RNA sequencing and bioinformatic analysis

Cardiac tissue was collected from wild-type and Ccl17-knockout mice 6 weeks after sham or TAC surgery (n=3 per group). Library preparation and sequencing were performed by OE Biotech (Shanghai, China). After assessment of RNA purity, concentration, and integrity, libraries were generated using the TruSeq Stranded mRNA LT Sample Prep Kit (Illumina) and sequenced as 150-bp paired-end reads on an Illumina HiSeq X Ten platform. Reads were quality-filtered using Trimmomatic, aligned to the mouse reference genome GRCm38.99 with HISAT2, and summarized at gene level using HTSeq-count. Differential expression was assessed using DESeq2, including version 1.22.2, with differentially expressed genes defined by an FDR<0.05 and |log□fold change|>1.5.^[25]^Principal component analysis, volcano plots, and row-Z-score-standardized heatmaps were used to visualize transcriptomic differences. Genes upregulated in Ccl17-knockout TAC hearts relative to wild-type TAC hearts were subjected to Reactome and WikiPathways over-representation analysis.

### Isolation and treatment of neonatal rat cardiac fibroblasts

Hearts from 1–2-day-old Sprague–Dawley rats were minced and subjected to serial digestion with 1 g/L type II collagenase at 37 °C. The resulting cell suspension was filtered through a cell strainer, and neonatal rat cardiac fibroblasts (NRCFs) were enriched by differential adhesion. Cells were seeded in six-well plates at 1 × 10⁶ cells per well and cultured in DMEM supplemented with 10% fetal bovine serum at 37 °C with 5% CO□. After serum starvation for 24 h, NRCFs were treated with vehicle, recombinant rat CCL17 (100 ng/mL; HY-P71902, MedChemExpress), CCL17 combined with the CCR4 antagonist AZD2098 (10 μM; HY-U00064, MedChemExpress), or TGF-β1 (10 ng/mL; HY-P7117, MedChemExpress) for 48 h. Cells and conditioned medium were then collected for RT–qPCR, immunoblotting, ELISA, and immunofluorescence analyses.

### siRNA transfection

Small interfering RNAs targeting rat Ccr4 or Egr1 and a non-targeting control siRNA were synthesized by Duona Pharmaceuticals. NRCFs were transfected at a final siRNA concentration of 10 nM using CALNP RNAi in vitro reagent according to the manufacturer’s instructions. Recombinant CCL17 (100 ng/mL) was added 24 h after transfection, and samples were collected 48 h later. Knockdown efficiency was verified by RT-qPCR. siRNA sequences are provided in Supplementary Table 3.

### Tissue and cellular immunofluorescence

Cardiac sections were fixed, subjected to antigen retrieval, permeabilized, blocked, and incubated overnight at 4 °C with antibodies against RFP and α-SMA. NRCFs were plated on coverslips coated with 1% gelatin, fixed after treatment with 4% paraformaldehyde for 15 min, permeabilized with 0.5% Triton X-100 for 5 min, blocked for 1 h, and incubated overnight at 4 °C with antibodies against IGFBP6 and α-SMA. Appropriate fluorescent secondary antibodies were applied the following day, and nuclei were counterstained with DAPI. Antibody details are provided in Supplementary Table 2. Images were acquired using a Leica SP8 laser-scanning confocal microscope with identical laser power, gain, exposure, and threshold settings across groups. Images were analysed using ImageJ.^[22]^Tissue RFP signal was expressed as background-corrected mean fluorescence intensity. Cellular IGFBP6 and α-SMA signals were quantified per cell or field and normalized to the control group.

### Promoter prediction and dual-luciferase assay

Putative transcription factor binding sites in the rat Igfbp6 promoter were predicted using the EGR1 binding motif (MA0162.5) from the JASPAR database.^[26]^EGR1 was selected based on motif prediction scores and transcription factor expression following CCL17 treatment. The wild-type Igfbp6 promoter and constructs carrying combined or individual mutations at sites S1–S3 were cloned into the pGL3-Basic vector. Mutations were introduced by replacing the corresponding binding sequences with adenine residues of equal length. Binding-site coordinates, wild-type sequences, and mutant sequences are provided in Supplementary Table 4

### Statistical analysis

Statistical analyses were performed using GraphPad Prism 9.4.1 and SPSS 26. Data are presented as mean±SEM, except for box-and-whisker plots, which show the median, interquartile range, and minimum and maximum values. Normality and homogeneity of variance were assessed using the Shapiro–Wilk and Levene tests, respectively. Two normally distributed groups with equal variances were compared using a two-sided unpaired Student t test; Welch t test was used when variances were unequal, and non-normally distributed data were analysed using the Mann–Whitney U test. Multiple groups were analysed by one-way or two-way analysis of variance, as appropriate, followed by Tukey multiple-comparison testing. Each point represents an independent participant, animal, or biological replicate; sample sizes are specified in the figure legends. Unless otherwise stated, two-sided P<0.05 was considered statistically significant.

## Results

### CCL17 is increased in patients with aortic stenosis and mice subjected to pressure overload

Because AS represents a clinically relevant form of chronic left ventricular pressure overload, and circulating proteomic signatures have been linked to myocardial remodelling in this setting,^[1, 2]^ we performed Olink proteomic profiling of serum samples from patients with AS and healthy controls (Figure 1A). Of the 70 samples initially profiled, 2 failed prespecified quality-control criteria and were excluded, leaving 34 participants in each group for subsequent analyses. The retained samples showed comparable normalized protein expression (NPX) distributions and high overall correlations, supporting the quality and consistency of the proteomic dataset (Figure S1A–S1E). Differential expression analysis identified 29 proteins associated with AS, including 11 upregulated and 18 downregulated proteins (Figure 1B and 1E). Among the upregulated proteins, serum CCL17 NPX levels were higher in patients with AS than in healthy controls (Figure 1C). Exploratory receiver operating characteristic analysis yielded an area under the curve of 0.748 (95% CI, 0.630–0.866; Figure 1D). CCL17 was therefore selected for further validation.

**Figure 1.**
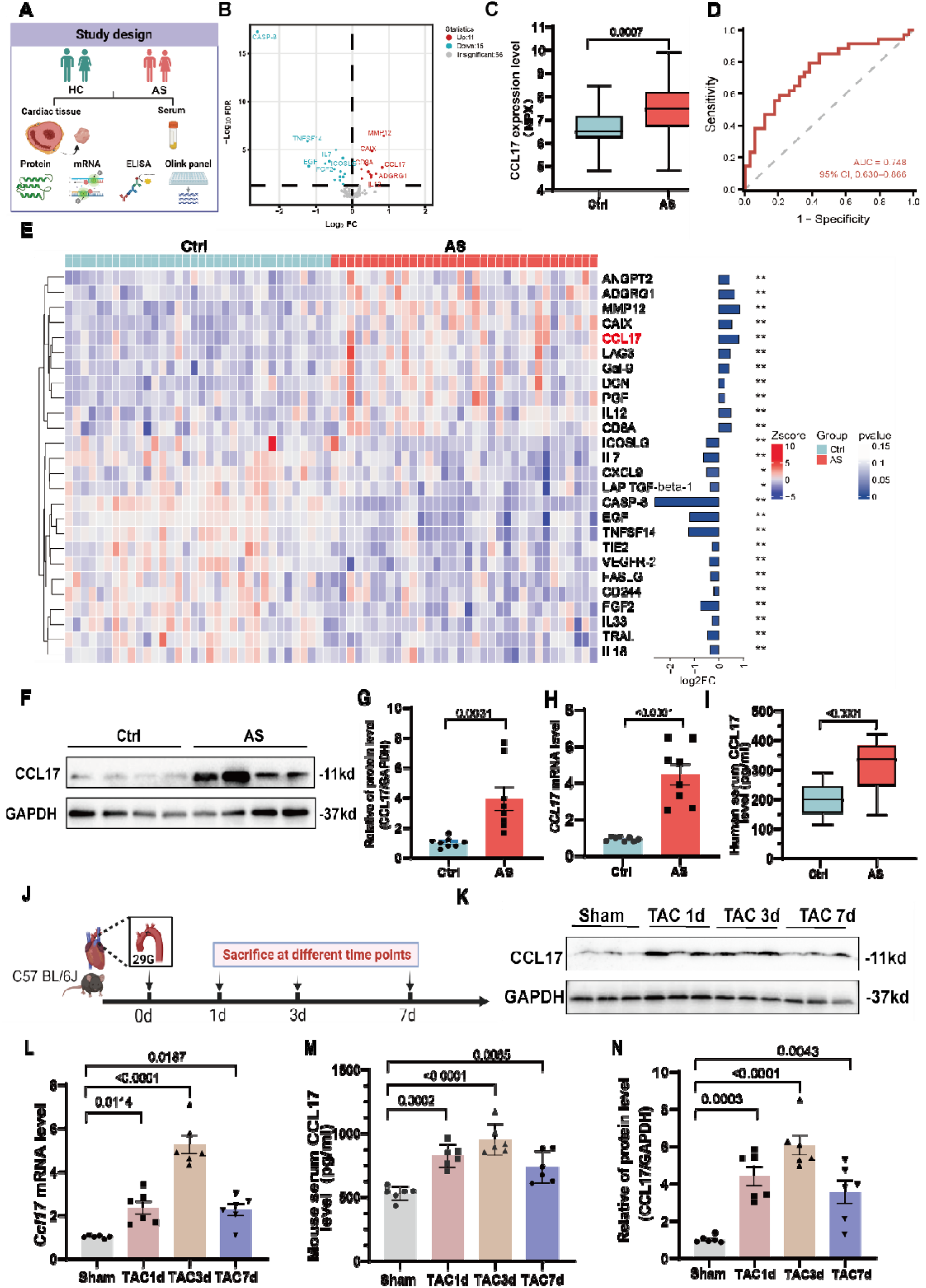
CCL17 is increased in patients with aortic stenosis and mice subjected to pressure overload. (A) Study design for Olink proteomic screening and subsequent validation in human serum and cardiac tissue. (B) Volcano plot of differentially expressed serum proteins in patients with aortic stenosis (AS) versus healthy controls (Ctrl). Red and blue indicate proteins upregulated and downregulated in AS, respectively; dashed lines indicate the prespecified significance thresholds. (C) Serum CCL17 normalized protein expression (NPX) in patients with AS and controls. (D) Receiver operating characteristic curve for serum CCL17 in discriminating patients with AS from controls. AUC, 0.748 (95% CI, 0.630–0.866). (E) Heatmap of the 26 differentially expressed serum proteins. Columns represent individual participants and rows represent proteins; colours indicate row-scaled NPX values. The accompanying bars show log2 fold changes, and CCL17 is highlighted in red. (F and G) Representative immunoblots (F) and quantification (G) of CCL17 protein in cardiac tissue from patients with AS and controls. GAPDH served as the loading control. (H) Relative cardiac *CCL17* mRNA expression in patients with AS and controls. (I) Serum CCL17 concentrations measured by enzyme-linked immunosorbent assay in patients with AS and controls. (J) Experimental timeline for transverse aortic constriction (TAC) and sample collection 1, 3, and 7 days after surgery. (K and N) Representative immunoblots (K) and quantification (N) of cardiac CCL17 protein after TAC. GAPDH served as the loading control. (L) Relative cardiac *Ccl17* mRNA expression after TAC. (M) Serum CCL17 concentrations after TAC. For (C and E), n=34 participants per group; for (F–H), n=8 participants per group; for (I), n=80 participants per group; and for (L–N), n=6 mice per group. Data are presented as mean±SEM, except for the box-and-whisker plots in (C and I), which show the median, interquartile range, and minimum and maximum values. Each symbol represents one participant or mouse. Comparisons between two groups were performed using a two-tailed unpaired Student t test (G and H) or Mann–Whitney U test (C and I). Time-course data were analysed using one-way ANOVA followed by Tukey’s multiple-comparison test. Exact P values are shown in the panels.

Consistent with the proteomic findings, immunoblotting demonstrated increased CCL17 protein abundance in cardiac tissues from patients with AS compared with controls (Figure 1F and 1G). Cardiac *CCL17* mRNA expression was similarly increased (Figure 1H). Enzyme-linked immunosorbent assay further confirmed higher serum CCL17 concentrations in patients with AS than in healthy controls (Figure 1I). Thus, the increase in circulating CCL17 identified by Olink profiling was corroborated in both serum and cardiac tissue from patients with AS.

We next examined whether CCL17 was similarly induced during experimental pressure overload. Wild-type mice were subjected to transverse aortic constriction (TAC), and cardiac tissue and serum were collected 1, 3, and 7 days after surgery (Figure 1J). Cardiac CCL17 protein abundance and *Ccl17* transcript levels increased by day 1, peaked at day 3, and remained elevated at day 7 after TAC compared with sham surgery (Figure 1K, 1L, and 1N). Serum CCL17 concentrations followed a similar temporal pattern (Figure 1M). This rapid induction was consistent with previous observations that cardiac *Ccl17* expression increases after ischaemic myocardial injury and angiotensin II-based cardiac stress,^[8, 9]^ and extended these findings to mechanical pressure overload. Together, these results identify CCL17 as a circulating and myocardial stress-responsive factor that is increased in human AS and rapidly induced after experimental pressure overload.

### CCL17 deficiency exacerbates pressure overload–induced cardiac dysfunction and adverse remodelling

Having established that CCL17 is increased in human AS and rapidly induced by experimental pressure overload, we next sought to determine whether this response promotes or restrains adverse cardiac remodelling. TAC is a well-established model of sustained left ventricular pressure overload that recapitulates the transition from compensatory hypertrophy to pathological remodelling and heart failure.^[21, 27]^ We therefore generated global *Ccl17*-knockout (*Ccl17*-KO) mice. Targeted deletion was confirmed by genotyping and by marked reductions in cardiac *Ccl17* expression and circulating CCL17 concentrations (Figure S2A–S2D).

WT and *Ccl17*-KO mice underwent TAC or sham surgery and were evaluated 6 weeks later (Figure 2A). Cardiac structural and functional indices were comparable between sham-operated WT and *Ccl17*-KO mice. After TAC, however, *Ccl17*-KO mice developed more severe left ventricular systolic dysfunction than WT mice, with lower ejection fraction and fractional shortening (Figure 2B, 2E, and 2F). CCL17 deficiency also augmented cardiac hypertrophy, as indicated by higher heart weight-to-tibia length and heart weight-to-body weight ratios and greater left ventricular mass (Figure 2C, 2D, and 2G).

**Figure 2.**
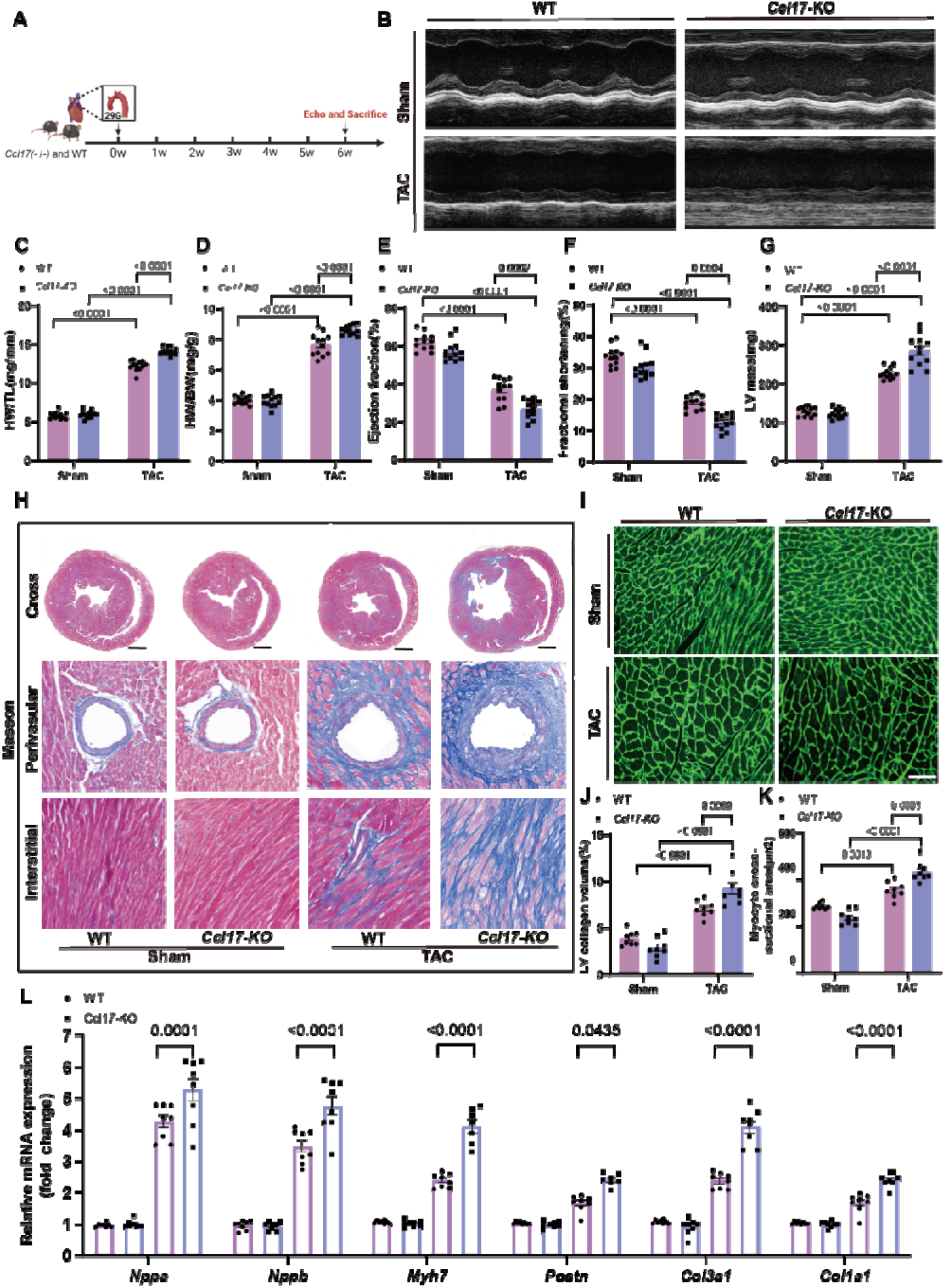
CCL17 deficiency exacerbates pressure overload-induced cardiac dysfunction and adverse remodelling. (A) Experimental design for transverse aortic constriction (TAC) or sham surgery in wild-type (WT) and *Ccl17*-knockout (*Ccl17*-KO) mice. Echocardiography was performed 4 and 6 weeks after surgery, and tissues were collected at 6 weeks. (B) Representative M-mode echocardiograms obtained 6 weeks after TAC or sham surgery. (C–G) Heart weight-to-tibia length ratio (C), heart weight-to-body weight ratio (D), left ventricular ejection fraction (E), fractional shortening (F), and left ventricular mass (G). (H) Representative Masson’s trichrome staining of whole-heart transverse sections and perivascular and interstitial myocardial regions. (I) Representative wheat germ agglutinin staining of cardiomyocyte membranes. (J and K) Quantification of left ventricular collagen volume fraction (J) and cardiomyocyte cross-sectional area (K). (L) Relative cardiac mRNA expression of the hypertrophic markers *Nppa*, *Nppb*, and *Myh7* and the fibroblast activation and fibrotic markers *Postn*, *Col3a1*, and *Col1a1*. Scale bars, 1 mm for whole-heart sections and 100 μm for higher-magnification Masson’s trichrome and wheat germ agglutinin images. For (C–G), n=12 mice per group; for (J–L), n=8 mice per group. Data are presented as mean±SEM. Each symbol represents one mouse. Statistical significance was assessed using two-way ANOVA followed by Tukey’s multiple-comparison test. Exact P values are shown in the panels.

Consistent with these functional and structural changes, *Ccl17*-KO-TAC hearts exhibited greater interstitial and perivascular collagen deposition and a higher left ventricular collagen volume fraction than WT-TAC hearts (Figure 2H and 2J). Cardiomyocyte cross-sectional area was also increased in *Ccl17*-KO-TAC mice (Figure 2I and 2K). Cardiomyocyte enlargement and fibroblast-driven extracellular matrix accumulation are cardinal components of maladaptive remodelling under sustained pressure overload.^[27, 28]^ Accordingly, cardiac expression of the hypertrophic markers *Nppa*, *Nppb*, and *Myh7* and the fibroblast activation and fibrotic markers *Postn*, *Col3a1*, and *Col1a1* was further increased after TAC in *Ccl17*-KO mice (Figure 2L).

Together, these loss-of-function findings indicate that endogenous CCL17 restrains pressure overload–induced cardiac hypertrophy, fibrosis, and systolic dysfunction. This protective effect contrasts with previous studies in myocardial infarction and angiotensin II–based models, in which genetic deletion or neutralization of CCL17 attenuated adverse cardiac remodelling.^[8, 9]^ These apparently divergent findings suggest that the function of CCL17 may depend on the nature and cellular context of cardiac injury. The unexpected worsening of cardiac remodelling after global *Ccl17* deletion prompted us to determine whether this protective effect could be attributed to myeloid-derived CCL17 and whether increasing CCL17 availability would confer reciprocal protection.

### AAV-mediated CCL17 augmentation attenuates pressure overload–induced cardiac remodelling

Because cells of the monocyte–macrophage lineage are a recognized source of CCL17, we next asked whether loss of CCL17 from Lyz2-expressing myeloid cells accounted for the adverse phenotype observed after global Ccl17 deletion. We generated Ccl17^fl/fl^;Lyz2^Cre/+^ mice and Cre-negative Ccl17^fl/fl^;Lyz2^+/+^ littermate controls (Figure S3A and S3B) and subjected both genotypes to sham or TAC surgery. Six weeks after TAC, myeloid-restricted Ccl17 deletion did not significantly alter heart weight-to-body weight or heart weight-to-tibia length ratios, left ventricular ejection fraction, or fractional shortening (Figure S3C–S3F). Thus, deletion of Ccl17 within the Lyz2 lineage was insufficient to reproduce the hypertrophic and functional phenotype observed after global Ccl17 deletion. Because this finding did not exclude a protective effect of increased CCL17 availability, we next examined whether macrophage-enriched CCL17 augmentation could protect the pressure-overloaded heart.

To address this question, we used an AAV-based gain-of-function strategy. AAV vectors enable sustained transgene expression in vivo, whereas F4/80 regulatory elements have been used to bias transgene expression towards cells of the monocyte–macrophage lineage.[29, 30] The AAV construct encoded Ccl17 under the control of the F4/80 promoter. Mice received AAV-F4/80-CCL17 or the corresponding AAV-F4/80-GFP control 1 week before TAC or sham surgery and were evaluated 6 weeks later (Figure 3A). Increased cardiac Ccl17 transcript abundance and serum CCL17 concentrations confirmed effective CCL17 augmentation (Figure S3G and S3H).

**Figure 3.**
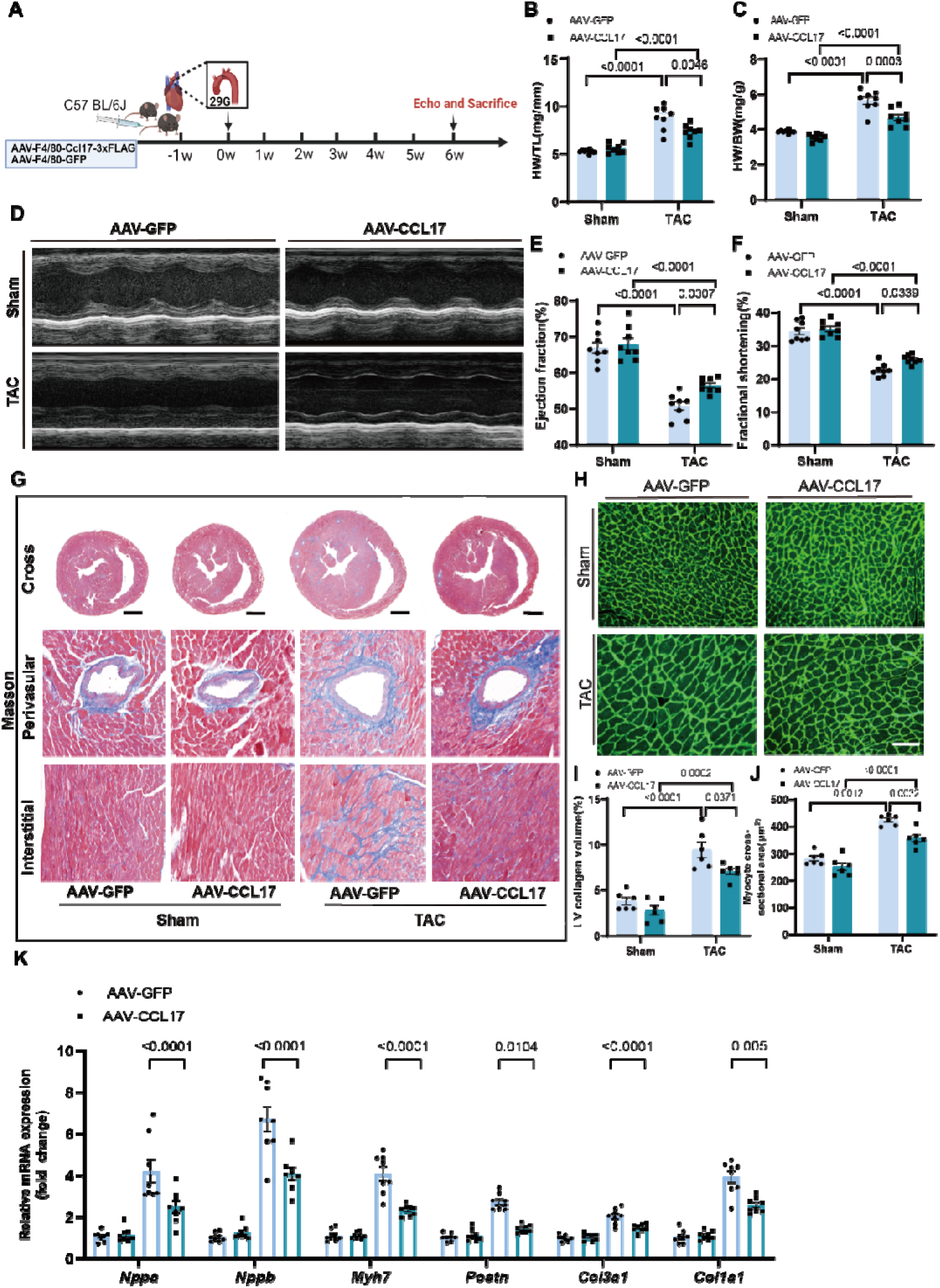
AAV-mediated CCL17 augmentation attenuates pressure overload-induced cardiac remodelling. (A) Experimental design. C57BL/6J mice received AAV-F4/80-*Ccl17*–3×FLAG (AAV-CCL17) or AAV-F4/80-GFP (AAV-GFP) 1 week before transverse aortic constriction (TAC) or sham surgery and were evaluated 6 weeks later. (B and C) Heart weight-to-tibia length ratio (B) and heart weight-to-body weight ratio (C). (D) Representative M-mode echocardiograms. (E and F) Left ventricular ejection fraction (E) and fractional shortening (F). (G) Representative Masson’s trichrome staining of whole-heart transverse sections and perivascular and interstitial myocardial regions. (H) Representative wheat germ agglutinin staining of cardiomyocyte membranes. (I and J) Quantification of left ventricular collagen volume fraction (I) and cardiomyocyte cross-sectional area (J). (K) Relative cardiac mRNA expression of the hypertrophic markers *Nppa*, *Nppb*, and *Myh7* and the fibroblast activation and fibrotic markers *Postn*, *Col3a1*, and *Col1a1*. Scale bars, 1 mm for whole-heart sections and 100 μm for higher-magnification Masson’s trichrome and wheat germ agglutinin images. For (B, C, E, F, and K), n=8 mice per group; for (I and J), n=6 mice per group. Data are presented as mean±SEM. Each symbol represents one mouse. Statistical significance was assessed using two-way ANOVA followed by Tukey’s multiple-comparison test. Exact P values are shown in the panels.

Compared with AAV-F4/80-GFP, AAV-F4/80-CCL17 preserved left ventricular systolic function after TAC, with higher ejection fraction and fractional shortening (Figure 3D–3F). AAV-F4/80-CCL17 also limited cardiac hypertrophy, as indicated by lower heart weight-to-tibia length and heart weight-to-body weight ratios and reduced left ventricular mass (Figure 3B and 3C; Figure S3C).

Histological analysis showed less interstitial and perivascular collagen deposition and a lower left ventricular collagen volume fraction in AAV-F4/80-CCL17-treated mice after TAC (Figure 3G and 3I). Cardiomyocyte cross-sectional area was also reduced (Figure 3H and 3J). Consistent with these changes, cardiac expression of the hypertrophic markers *Nppa*, *Nppb*, and *Myh7* and the fibroblast activation and fibrotic markers *Postn*, *Col3a1*, and *Col1a1* was lower following AAV-F4/80-CCL17 treatment (Figure 3K).

Together, these gain-of-function findings complement the global knockout data and establish that increasing CCL17 availability attenuates pressure overload–induced cardiac dysfunction and adverse remodelling. The reciprocal effects observed with CCL17 deletion and augmentation in the present study support a protective role for CCL17 in the context of TAC-induced pressure overload.

### CCL17 restrains IGFBP6 induction in pressure-overloaded hearts

To identify molecular programmes associated with the cardioprotective effects of CCL17, we performed bulk RNA sequencing of cardiac tissues from WT and *Ccl17*-KO mice 6 weeks after sham or TAC surgery. Principal component analysis showed distinct clustering of the four experimental groups (Figure 4A). Across the indicated comparisons, TAC induced extensive transcriptional remodelling, whereas direct comparison of *Ccl17*-KO-TAC and WT-TAC hearts identified 258 upregulated and 190 downregulated genes (Figure 4B and 4C).

**Figure 4.**
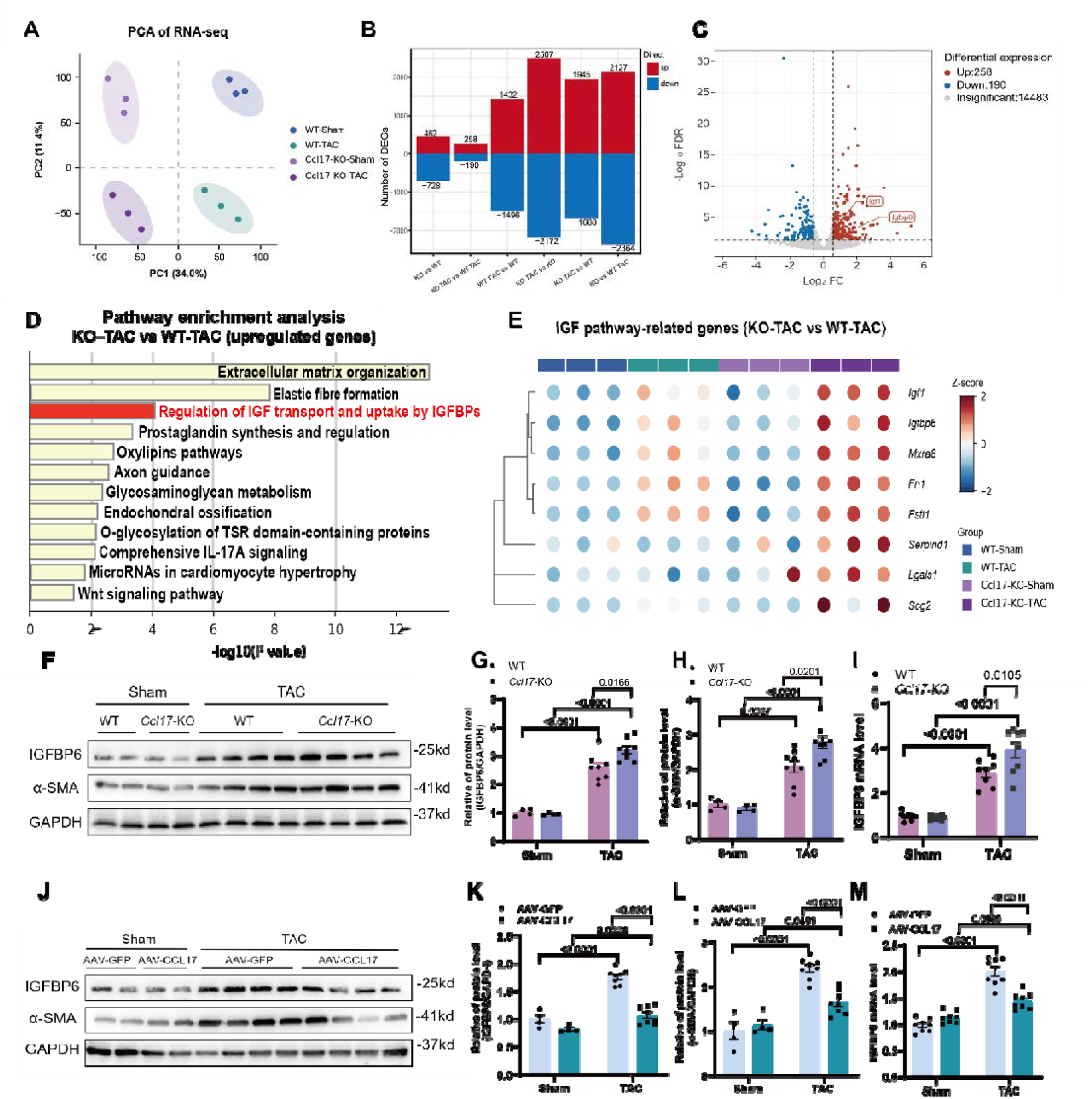
CCL17 restrains IGFBP6 induction in pressure-overloaded hearts. (A) Principal component analysis of cardiac RNA-sequencing profiles from wild-type (WT) and *Ccl17*-knockout (*Ccl17*-KO) mice after sham or transverse aortic constriction (TAC) surgery. (B) Numbers of upregulated and downregulated genes across the indicated pairwise comparisons. (C) Volcano plot of differentially expressed genes in *Ccl17*-KO-TAC versus WT-TAC hearts, showing 258 upregulated and 190 downregulated genes. *Igf1* and *Igfbp6* are indicated. Differentially expressed genes were defined by a false discovery rate (FDR) <0.05 and an absolute log2 fold change >1.5. (D) Over-representation analysis of genes upregulated in *Ccl17*-KO-TAC versus WT-TAC hearts using Reactome and WikiPathways terms. The Reactome term ‘Regulation of IGF transport and uptake by IGFBPs’ is highlighted. (E) Heatmap of the eight differentially expressed genes assigned to the IGFBP-associated Reactome term. Columns represent individual mice and rows represent genes; expression values were standardized as row Z scores. (F) Representative immunoblots of cardiac IGFBP6 and α-smooth muscle actin (α-SMA) in WT and *Ccl17*-KO mice after sham or TAC surgery. GAPDH served as the loading control. (G and H) Quantification of IGFBP6 (G) and α-SMA (H) protein abundance. (I) Relative cardiac *Igfbp6* mRNA expression. (J) Representative immunoblots of cardiac IGFBP6 and α-SMA in mice treated with AAV-F4/80-GFP or AAV-F4/80-CCL17 after sham or TAC surgery. GAPDH served as the loading control. (K and L) Quantification of IGFBP6 (K) and α-SMA (L) protein abundance. (M) Relative cardiac *Igfbp6* mRNA expression. For (A–E), n=3 mice per group; for (G–I), n=8 mice per group; and for (K–M), n=4 mice per sham group and n=8 mice per TAC group. Data are presented as mean±SEM. Each symbol represents one mouse. Statistical significance was assessed using two-way ANOVA followed by Tukey’s multiple-comparison test. Exact P values are shown in the panels.

Pathway enrichment analysis of genes upregulated in *Ccl17*-KO-TAC hearts revealed extracellular matrix organization and elastic fibre formation, together with regulation of insulin-like growth factor transport and uptake by insulin-like growth factor-binding proteins (IGFBPs) (Figure 4D). IGFBPs regulate the stability, tissue availability and receptor accessibility of IGFs and can also exert IGF-independent biological effects.^[15]^ Both *Igf1* and *Igfbp6* were upregulated in *Ccl17*-KO-TAC hearts, and the eight differentially expressed genes assigned to the IGFBP-associated pathway exhibited a distinct expression pattern in this group (Figure 4C and 4E).

We next examined these eight candidates in two reciprocal models of CCL17 loss and augmentation. We next examined the IGFBP pathway-associated candidates in two reciprocal models of CCL17 loss and augmentation. Except for *Igfbp6*, the remaining candidate genes did not show concordant CCL17-dependent changes across the two models (Figure S4A and S4B). Notably, *Igf1* transcript abundance was further increased in *Ccl17*-KO hearts after TAC compared with WT-TAC hearts, but this change was not reciprocally reproduced following AAV-mediated CCL17 augmentation (Figure S4A and S4B). Moreover, although TAC increased cardiac IGF1 protein abundance, IGF1 protein levels did not differ between WT-TAC and *Ccl17*-KO-TAC hearts, indicating a lack of concordance between the transcript-level and protein-level findings (Figure S4C). The consistent reciprocal regulation of *Igfbp6*, together with the absence of a corresponding IGF1 protein change, therefore led us to prioritize IGFBP6 for subsequent mechanistic investigation.Consistent with the transcriptomic findings, TAC-induced increases in cardiac IGFBP6 protein and *Igfbp6* mRNA were further enhanced in *Ccl17*-KO mice compared with WT mice and were accompanied by increased α-SMA abundance (Figure 4F–4I). Conversely, AAV-F4/80-CCL17 attenuated the TAC-induced increases in IGFBP6 and α-SMA protein abundance and reduced cardiac *Igfbp6* expression compared with AAV-F4/80-GFP (Figure 4J–4M). These reciprocal findings prioritized IGFBP6 as a CCL17-responsive candidate associated with pressure overload–induced fibrotic remodelling, consistent with recent evidence identifying fibroblast-derived IGFBP6 as a promoter of myofibroblast activation and adverse cardiac remodelling.^[16]^

### CCL17 suppresses IGFBP6 expression in cardiac fibroblasts through CCR4–EGR1 signalling

Having identified IGFBP6 as a CCL17-responsive candidate, we next examined the cellular context and signalling mechanism linking CCL17 to IGFBP6. In *Ccl17*-tdTomato reporter mice, cardiac reporter activity was low under basal conditions but markedly increased 3 days after TAC and localized within α-SMA-rich remodelling regions (Figure 5A–5C). Reporter validation and genotyping are shown in Figure S5A and Figure S5B

**Figure 5.**
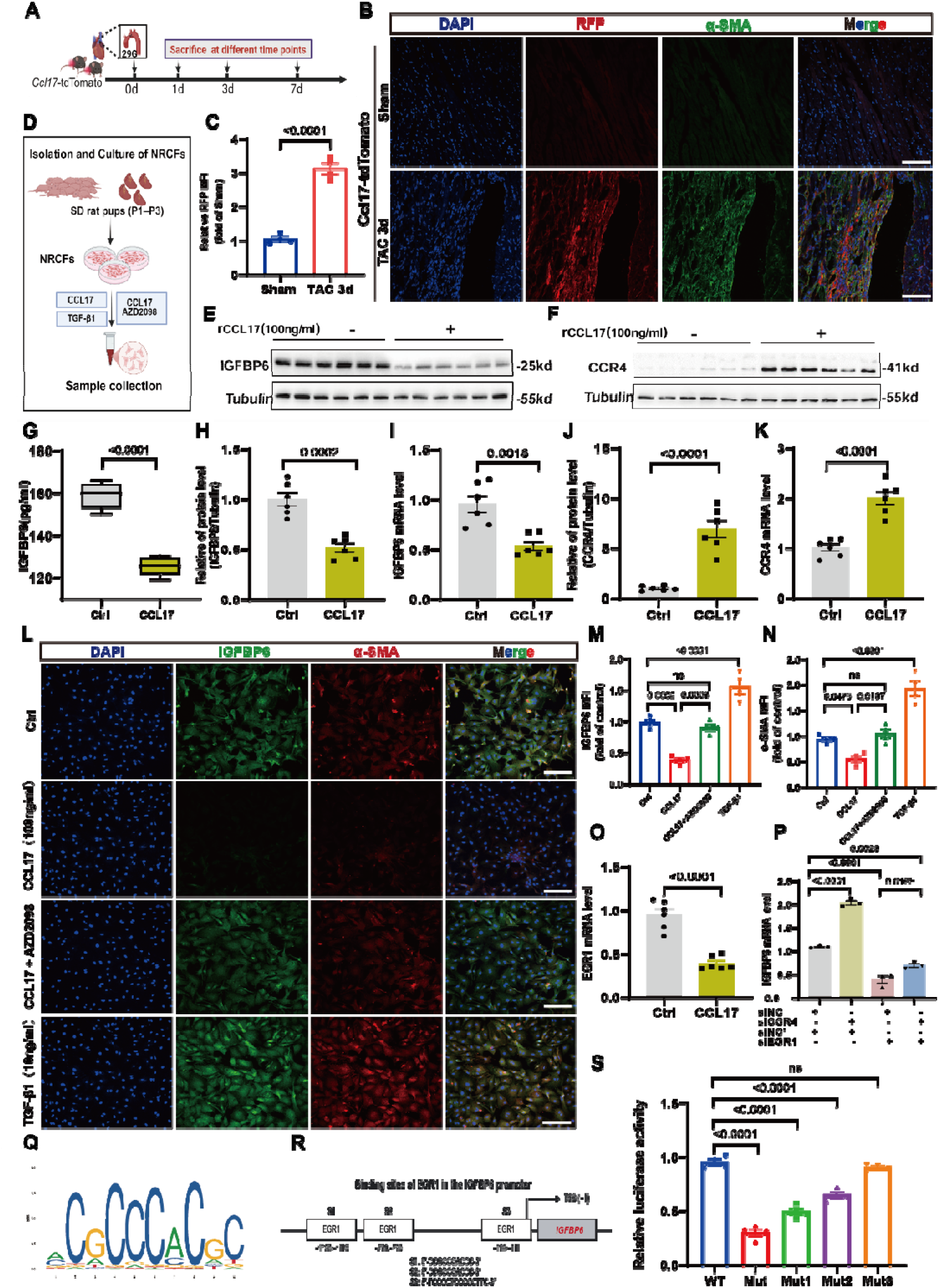
CCL17 suppresses IGFBP6 expression and myofibroblast activation through CCR4–EGR1 signalling. (A) Experimental design for temporal analysis of cardiac CCL17 expression in *Ccl17*-tdTomato reporter mice subjected to transverse aortic constriction (TAC). Hearts were collected before surgery and 1, 3, and 7 days after TAC. (B) Representative immunofluorescence images of DAPI, RFP, α-smooth muscle actin (α-SMA), and merged signals in cardiac sections from sham-operated and TAC-treated *Ccl17*-tdTomato mice at day 3. (C) Background-corrected RFP mean fluorescence intensity (MFI) in cardiac sections from sham-operated and TAC-treated *Ccl17*-tdTomato mice at day 3, expressed relative to sham. (D) Schematic of neonatal rat cardiac fibroblast (NRCF) isolation, culture, treatment, and sample collection. (E and F) Representative immunoblots of IGFBP6 (E) and CCR4 (F) in NRCFs treated with vehicle or recombinant CCL17 (rCCL17; 100 ng/mL). Tubulin served as the loading control. (G) IGFBP6 concentrations in conditioned medium from vehicle-and rCCL17-treated NRCFs, measured by enzyme-linked immunosorbent assay. (H and I) Quantification of cellular IGFBP6 protein abundance (H) and relative *Igfbp6* mRNA expression (I) after rCCL17 treatment. (J and K) Quantification of CCR4 protein abundance (J) and relative *Ccr4* mRNA expression (K) after rCCL17 treatment. (L) Representative immunofluorescence images of DAPI, IGFBP6, α-SMA, and merged signals in NRCFs treated with vehicle, rCCL17, rCCL17 plus the CCR4 antagonist AZD2098, or transforming growth factor-β1 (TGF-β1). (M and N) Quantification of IGFBP6 (M) and α-SMA (N) MFI in the indicated treatment groups, expressed relative to control. (O) Relative *Egr1* mRNA expression in NRCFs treated with vehicle or rCCL17. (P) Relative *Igfbp6* mRNA expression in rCCL17-treated NRCFs transfected with control small interfering RNA (siNC), CCR4-targeting siRNA (si*Ccr4*), EGR1-targeting siRNA (si*Egr1*), or the indicated combination of si*Ccr4* and si*Egr1*. (Q) Predicted EGR1 DNA-binding motif. (R) Schematic of three predicted EGR1-responsive elements (S1–S3) within the rat *Igfbp6* promoter; positions are indicated relative to the transcription start site. (S) Dual-luciferase reporter analysis of the wild-type *Igfbp6* promoter construct (WT) and constructs carrying combined or individual mutations in the predicted EGR1-responsive elements (Mut and Mut1–Mut3). Luciferase activity is expressed relative to the WT construct. Scale bars, 100 μm. For C, n=4 mice per group. Data are presented as mean±SEM, except for the box-and-whisker plot in (G), which shows the median, interquartile range, and minimum and maximum values. Each symbol represents one mouse in (C) or one independent biological replicate in the cell experiments. Comparisons between two groups were performed using two-tailed unpaired Student t tests (C, G–K, and O). Comparisons among multiple groups were performed using one-way ANOVA followed by Tukey’s multiple-comparison test (M, N, and S) or two-way ANOVA followed by Tukey’s multiple-comparison test (P). Exact P values are shown in the panels.

We subsequently isolated neonatal rat cardiac fibroblasts (NRCFs) for subsequent mechanistic studies (Figure 5D). Recombinant CCL17 reduced IGFBP6 concentrations in conditioned medium (Figure 5G), together with decreases in cellular IGFBP6 protein and *Igfbp6* transcript abundance (Figure 5E, 5H, and 5I). In contrast, CCL17 increased CCR4 protein and *Ccr4* transcript expression (Figure 5F, 5J, and 5K). Given that CCR4 is an established functional receptor for CCL17,^[31]^ we next examined whether CCR4 mediated these effects. AZD2098 is a potent and bioavailable small-molecule CCR4 antagonist.^[32]^

Immunofluorescence analysis further showed that CCL17 reduced IGFBP6 and α-SMA abundance in NRCFs. These effects were reversed by the CCR4 antagonist AZD2098, whereas TGF-β1 increased both IGFBP6 and α-SMA as a positive control for fibroblast activation (Figure 5L–5N). These findings indicated that pharmacological blockade of CCR4 counteracted the effects of CCL17 on IGFBP6 expression and fibroblast activation.

To identify candidate transcriptional regulators that might link CCR4 signalling to *Igfbp6* expression, we screened the predicted rat *Igfbp6* promoter sequence using the JASPAR transcription factor–binding profile database. This analysis identified three high-scoring putative EGR1-responsive elements within the *Igfbp6* promoter (Figure 5Q and 5R). Subsequent qPCR screening in CCL17-treated NRCFs showed that *Egr1* expression was reduced, prompting us to prioritize EGR1 for further investigation (Figure 5O; Figure S5C-S5E). This selection was also supported by previous evidence implicating EGR1-dependent transcription in cardiac fibroblast activation and fibrotic remodelling.^[33]^

Silencing *Ccr4* increased *Igfbp6* expression, whereas silencing *Egr1* reduced *Igfbp6* expression and blunted its induction following *Ccr4* knockdown (Figure 5P). The efficiencies of *Ccr4* and *Egr1* knockdown were confirmed at the transcript and protein levels (Figure S5F and S5G). In dual-luciferase reporter assays, combined or individual mutation of the predicted EGR1-responsive elements differentially reduced *Igfbp6* promoter activity, with the most pronounced reduction observed following combined mutation (Figure 5S).Collectively, these findings are consistent with a model in which CCL17–CCR4 signalling restrains *Igfbp6* expression through EGR1-dependent transcriptional regulation.

### Fibroblast-directed *Igfbp6* silencing attenuates adverse remodelling caused by CCL17 deficiency

To determine whether increased IGFBP6 functionally contributes to the adverse cardiac remodelling associated with CCL17 deficiency, we performed a fibroblast-directed rescue experiment. This approach was supported by recent evidence identifying fibroblast-derived IGFBP6 as a profibrotic mediator of cardiac remodelling.^[16]^ *Ccl17*-KO mice received AAV9-COL1A2-sh*Igfbp6* or the corresponding AAV9-COL1A2-shCtrl 1 week before TAC, whereas WT mice receiving AAV9-COL1A2-shCtrl served as the reference group. Cardiac structure and function were evaluated 6 weeks after TAC (Figure 6A).

**Figure 6.**
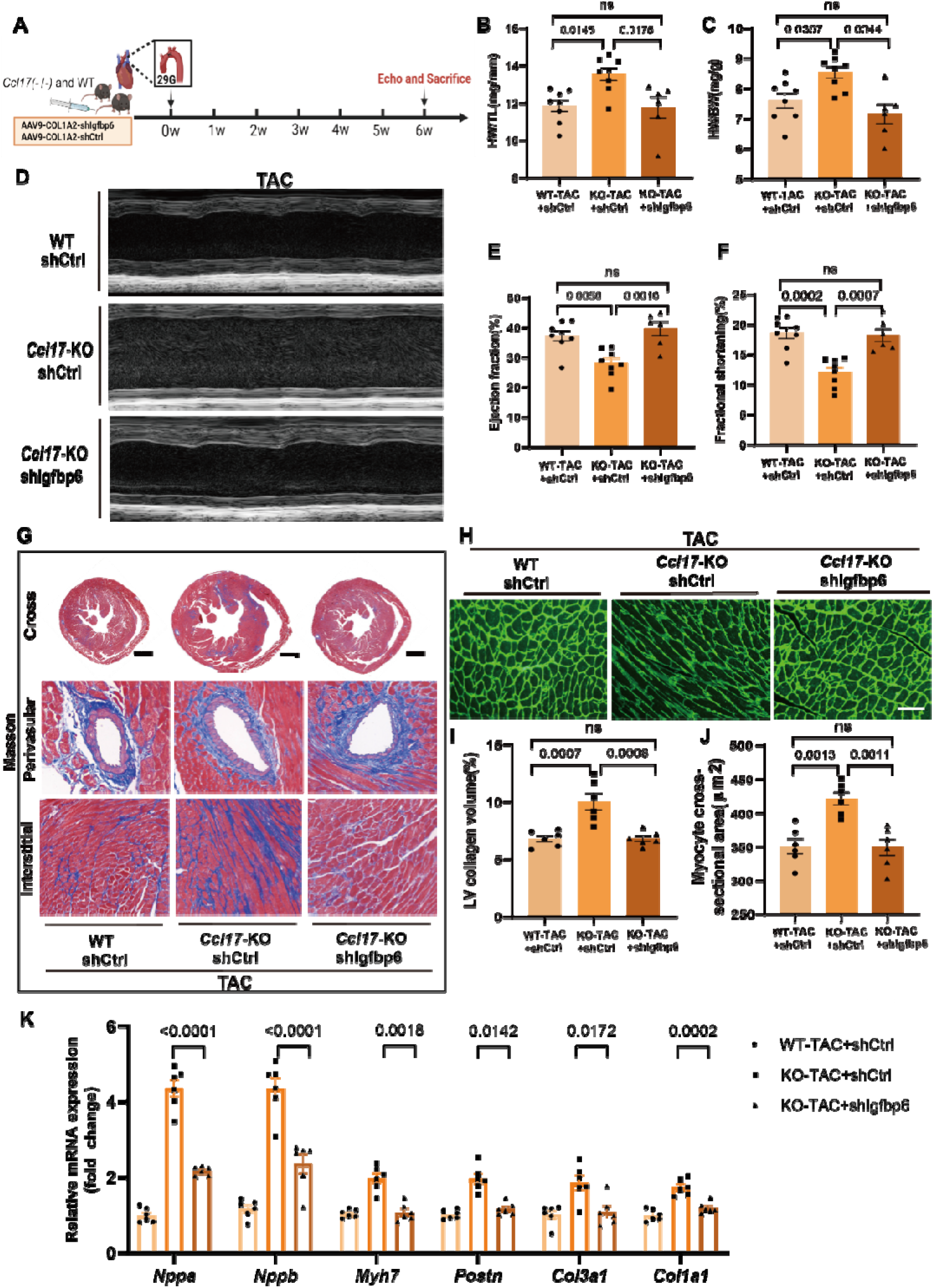
AAV9-COL1A2-mediated *Igfbp6* silencing attenuates adverse cardiac remodelling in *Ccl17*-deficient mice. (A) Experimental protocol for fibroblast-directed *Igfbp6* silencing and transverse aortic constriction (TAC). Wild-type (WT) mice received AAV9-COL1A2-shCtrl, whereas *Ccl17*-knockout (*Ccl17*-KO) mice received AAV9-COL1A2-shCtrl or AAV9-COL1A2-sh*Igfbp6* 1 week before TAC. Echocardiography and tissue collection were performed 6 weeks after TAC. (B and C) Heart weight-to-tibia length ratio (B) and heart weight-to-body weight ratio (C). (D–F) Representative M-mode echocardiograms (D) and quantification of left ventricular ejection fraction (E) and fractional shortening (F). (G) Representative Masson’s trichrome staining of whole-heart transverse sections and perivascular and interstitial myocardial regions. (H) Representative wheat germ agglutinin staining of cardiomyocyte membranes. (I and J) Quantification of left ventricular collagen volume fraction (I) and cardiomyocyte cross-sectional area (J). (K) Relative cardiac mRNA expression of the hypertrophic markers *Nppa*, *Nppb*, and *Myh7* and the fibroblast activation and fibrotic markers *Postn*, *Col3a1*, and *Col1a1*. Scale bars, 1 mm for whole-heart sections and 100 μm for higher-magnification Masson’s trichrome and wheat germ agglutinin images. For (B-F), n=8 mice per group; for (I-K), n=6 mice per group Data are presented as mean±SEM. Each symbol represents one mouse. Statistical significance was assessed using one-way ANOVA followed by Tukey’s multiple-comparison test. Exact P values are shown in the panels.

Compared with WT-TAC mice receiving shCtrl, *Ccl17*-KO-TAC mice receiving shCtrl exhibited greater cardiac hypertrophy, as reflected by increased heart weight-to-tibia length and heart weight-to-body weight ratios (Figure 6B and 6C). They also developed more severe left ventricular systolic dysfunction, with lower ejection fraction and fractional shortening (Figure 6D–6F). Fibroblast-directed *Igfbp6* silencing attenuated cardiac hypertrophy and improved systolic function, restoring these parameters towards those observed in WT-TAC controls (Figure 6B–6F).

Histological analysis showed greater interstitial and perivascular collagen deposition in *Ccl17*-KO-TAC mice receiving shCtrl than in WT-TAC controls. These changes were attenuated by *Igfbp6* silencing, as demonstrated by reduced collagen deposition and a lower left ventricular collagen volume fraction (Figure 6G and 6I). Cardiomyocyte cross-sectional area was also reduced following *Igfbp6* silencing (Figure 6H and 6J).

Consistent with these functional and histological improvements, *Igfbp6* silencing reduced cardiac expression of the hypertrophic markers *Nppa, Nppb*, and *Myh7*, as well as the fibroblast activation and fibrotic markers *Postn*, *Col3a1*, and *Col1a1*, in *Ccl17*-KO-TAC mice (Figure 6K). Collectively, these rescue experiments identify increased IGFBP6 as a functionally important downstream mediator of the cardiac dysfunction, hypertrophy, and fibrosis associated with CCL17 deficiency. Together with the preceding loss-and gain-of-function studies, these findings support a protective mechanism in which CCL17–CCR4 signalling restrains EGR1-dependent IGFBP6 expression during pressure overload.

## Discussion

The present study identifies a previously unrecognized protective role for CCL17 during sustained pressure overload. Circulating and myocardial CCL17 abundance was increased in patients with aortic stenosis and was rapidly induced after TAC in mice. Despite this increase, genetic deletion of *Ccl17* aggravated ventricular dysfunction, hypertrophy and fibrosis, whereas AAV-mediated CCL17 augmentation produced the reciprocal phenotype. Transcriptomic and mechanistic analyses further identified IGFBP6 as a CCL17-responsive, fibroblast-associated mediator, and fibroblast-directed *Igfbp6* silencing partially rescued the adverse phenotype caused by CCL17 deficiency. Collectively, these findings support a model in which CCL17 restrains pressure overload-induced remodelling, at least in part by limiting CCR4–EGR1-associated *Igfbp6* transcription in cardiac fibroblasts. More broadly, they suggest that an increase in a circulating inflammatory mediator during cardiovascular disease does not necessarily indicate a pathogenic response, but may instead reflect an endogenous attempt to preserve tissue homeostasis.

Chemokines are rapidly induced after cardiac injury, where they coordinate immune-cell recruitment and immune-stromal interactions that shape tissue repair^[34, 35]^. In our TAC model, circulating and myocardial CCL17 levels increased by day 1 and peaked on day 3, preceding the assessment of ventricular remodelling at six weeks. This temporal pattern, together with the reciprocal effects observed in loss-and gain-of-function experiments, identifies CCL17 as an early stress-responsive chemokine with a protective role during sustained pressure overload. CCL17 has also been reported to exert protective effects in viral myocarditis by limiting regulatory T-cell recruitment^[36]^. By contrast, previous studies of myocardial infarction and ageing-or angiotensin II-associated cardiac remodelling implicated CCL17 in detrimental effects mediated through immune-cell regulation ^[8, 9]^. These divergent findings suggest that the effects of CCL17 may vary with the nature of cardiac stress and the cellular programmes engaged. This context dependence raises the possibility that CCL17 acts on non-immune cells involved in myocardial remodelling during pressure overload.

Cardiac fibroblasts are plausible targets because they integrate mechanical and inflammatory cues to regulate extracellular matrix deposition and cardiac remodelling ^[4, 5, 37–39]^. Following TAC, *Ccl17* reporter activity increased in α-SMA-rich myocardial regions, spatially associating the induced chemokine response with areas of active remodelling. In isolated cardiac fibroblasts, recombinant CCL17 reduced IGFBP6 expression and α-SMA abundance, whereas pharmacological blockade of CCR4 counteracted these effects. The increase in *Igfbp6* expression after *Ccr4* silencing further implicated CCR4 in this response.However, recent evidence identifying CCR8 as an alternative receptor for CCL17 underscores the complexity of its receptor interactions^[14]^. Notably, CCL17-CCR4 signalling has been reported to promote fibroblast activation in pulmonary fibrosis^[40]^, suggesting that stromal responses to CCL17 may vary across tissue contexts.Together, these findings extend the biological role of CCL17 beyond immune-cell trafficking by demonstrating direct fibroblast responsiveness and implicating CCR4-associated signalling in its protective effects during pressure overload.

Growth factor signalling is regulated not only by ligand abundance but also by proteins that control ligand availability and cellular responsiveness. The insulin-like growth factor-binding proteins (IGFBPs) constitute one such regulatory layer, modulating IGF bioavailability while also exerting IGF-independent effects ^[15]^. In our study, although *Igf1* and *Igfbp6* expression were both increased in *Ccl17*-deficient hearts after TAC, IGF1 protein did not change consistently across reciprocal CCL17 interventions. By contrast, IGFBP6 was increased by CCL17 deficiency and suppressed by CCL17 augmentation, indicating that this regulatory layer is particularly responsive to CCL17. Importantly, fibroblast-targeted *Igfbp6* silencing attenuated cardiac hypertrophy, fibrosis, and systolic dysfunction in *Ccl17*-deficient mice, establishing a functional contribution of IGFBP6 to the adverse remodelling phenotype in this context. Together with previous evidence implicating fibroblast-derived IGFBP6 in cardiac fibrosis ^[16]^, these findings connect chemokine signalling with growth factor-associated fibroblast programmes and implicate suppression of IGFBP6 as one mechanism through which CCL17 restrains adverse remodelling.

A key question arising from this chemokine-growth-regulatory connection is how extracellular CCL17 signalling is translated into changes in fibroblast gene expression. Integration of *Igfbp6* promoter analysis with transcription-factor expression screening identified EGR1, a stress-responsive transcription factor implicated in cardiac fibroblast activation and fibrosis ^[33]^, as a candidate regulator. Consistent with this prediction, CCL17 reduced *Egr1* expression, whereas *Egr1* silencing attenuated the increase in *Igfbp6* expression induced by *Ccr4* knockdown. Mutation of predicted EGR1-responsive elements within the *Igfbp6* promoter further reduced reporter activity. Together, these results support a model in which CCL17-CCR4 signalling suppresses EGR1-associated transcriptional regulation of *Igfbp6*. Cheng et al. previously reported that intracellular IGFBP6 interacts with EGR1 to promote Mfap4 transcription ^[16]^. Whereas their findings position IGFBP6 as a modulator of EGR1-dependent transcription, our results place EGR1 upstream of *Igfbp6* expression, revealing a distinct regulatory relationship between these molecules in cardiac fibroblasts.

The translational relevance of these findings is suggested by the concordant elevation of circulating and myocardial CCL17 in patients with AS and its rapid induction after TAC in mice. This pattern highlights an important consideration when interpreting circulating inflammatory proteins: increased abundance in cardiovascular disease may represent an adaptive response rather than simply a pathogenic signal. In AS, determining whether CCL17 reflects haemodynamic burden, myocardial adaptation, or treatment-related changes may therefore be more informative than evaluating its diagnostic performance alone. Longitudinal studies incorporating serial CCL17 measurements with assessments of myocardial remodelling, ventricular function, and clinical outcomes before and after aortic valve replacement will be needed to clarify its clinical significance and determine whether the protective pathway identified in experimental pressure overload is relevant to human disease.^[41, 42]^

Several limitations warrant consideration. First, although CCL17 directly modulated isolated cardiac fibroblasts through CCR4-associated signalling, its endogenous cellular sources and principal responding cell populations in vivo remain unresolved^[43]^. Global *Ccl17* deletion, F4/80-driven CCL17 augmentation, and spatial reporter analyses cannot definitively establish this source-target relationship, while COL1A2-directed *Igfbp6* silencing may not be exclusively fibroblast specific. Second, bulk cardiac transcriptomics cannot distinguish cell-intrinsic changes in gene expression from shifts in cellular composition. Moreover, the genetic perturbation and promoter-reporter experiments support an EGR1-associated mechanism but do not demonstrate endogenous EGR1 occupancy at the Igfbp6 locus or define the receptor-proximal signals linking CCR4 to EGR1. Third, although Igfbp6 silencing established a functional contribution to the phenotype, it does not exclude the involvement of other CCL17-responsive pathways, and the relative contributions of IGF-dependent and IGF-independent mechanisms remain unclear. Finally, the human observations were cross-sectional, and the exploratory diagnostic analysis requires independent validation. Neither provides evidence that CCL17 is protective in patients. Cell-specific genetic studies and longitudinal human investigations will be required to resolve these mechanisms and establish their clinical significance.

In conclusion, our study identifies CCL17 as an endogenous protective signal that limits adverse cardiac remodelling during sustained pressure overload, contrasting with its previously reported detrimental effects in other cardiac injury settings. We demonstrate that cardiac fibroblasts respond directly to CCL17 and identify CCR4-EGR1-associated suppression of IGFBP6 as one functionally relevant mechanism, thereby extending the biological role of CCL17 beyond immune-cell trafficking. More broadly, this work reveals a protective dimension of chemokine-stromal communication and shows how immune-associated signals can modulate growth-regulatory programmes in cardiac fibroblasts during myocardial stress.

## Acknowledgements

The authors thank all participants and tissue donors who contributed samples to this study. The authors also thank LC-Bio Technology Co., Ltd. for technical support with the proteomic analysis.

## Data Availability

All data are available in the manuscript or the supplementary materials.

