## Supplementary Figures and Tables for "CCL17-CCR4 Signalling Attenuates Pressure Overload-Induced Cardiac Remodelling Through IGFBP6 Suppression in Cardiac Fibroblasts"

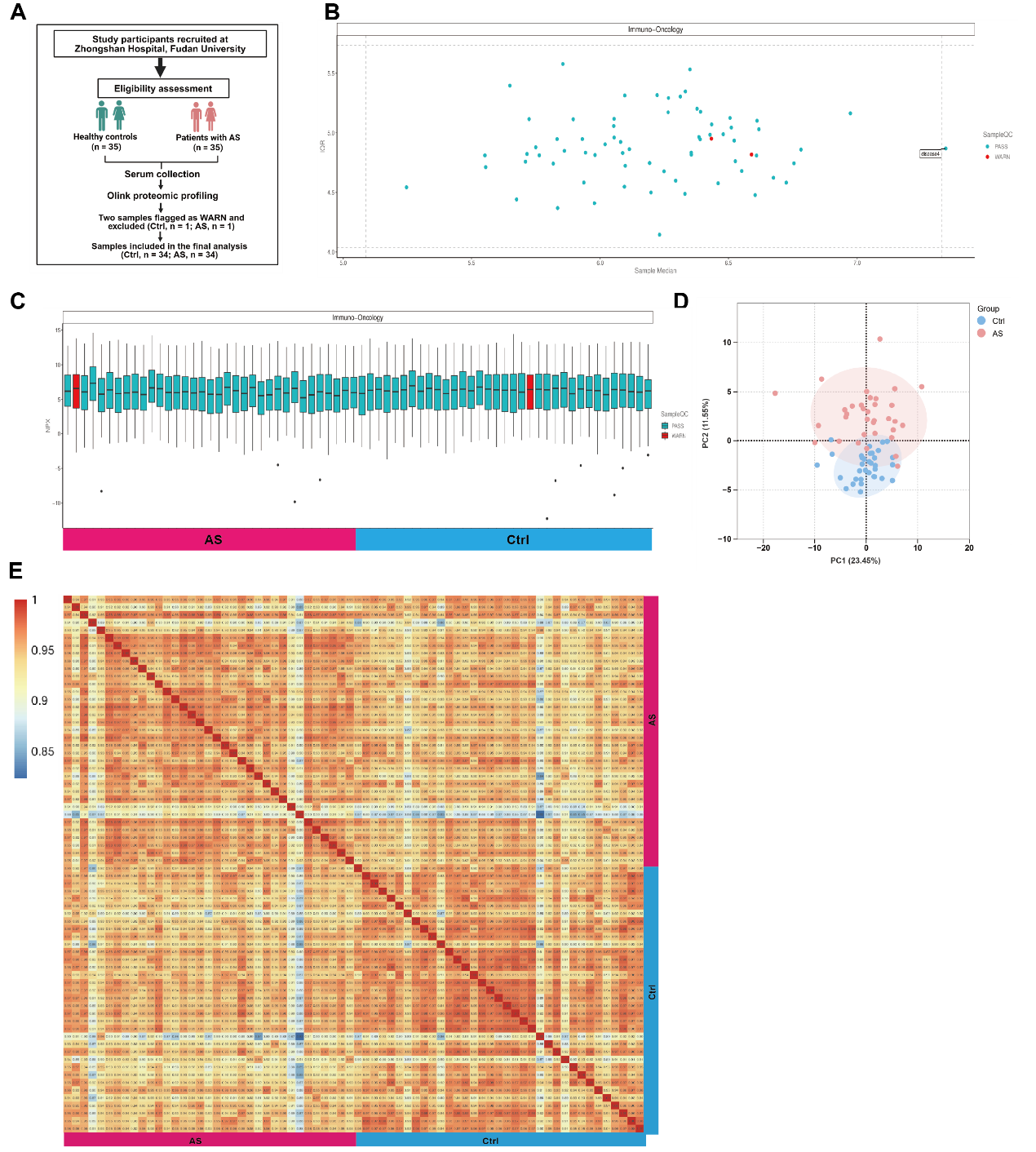

**Supplementary Figure 1. Study workflow and quality assessment of the Olink proteomic dataset.**

(A) Flow diagram of participant recruitment, serum collection, Olink proteomic profiling, quality-control assessment, and sample inclusion. Of the 70 serum samples initially profiled (Ctrl, n=35; AS, n=35), 2 samples flagged as WARN were excluded (Ctrl, n=1; AS, n=1), leaving 34 samples per group for the final analysis. (B) Sample-level quality-control assessment based on the median and interquartile range of NPX values. Samples classified as PASS and WARN are shown in turquoise and red, respectively; dashed lines indicate the quality-control boundaries. (C) NPX distributions across the profiled serum samples. Box plots show the distribution of all measured proteins within each sample, with WARN samples highlighted in red. The colored bar below indicates the clinical group. (D) Principal component analysis of the serum proteomic profiles retained after quality control. Each symbol represents 1 participant, and shaded areas indicate 95% confidence ellipses (Ctrl, n=34; AS, n=34). The percentages of variance explained by principal components 1 and 2 are shown on the axes. (E) Sample-to-sample Spearman correlation heatmap based on all measured proteins in the quality-controlled dataset. Rows and columns represent individual participants, and colors indicate Spearman correlation coefficients (Ctrl, n=34; AS, n=34).

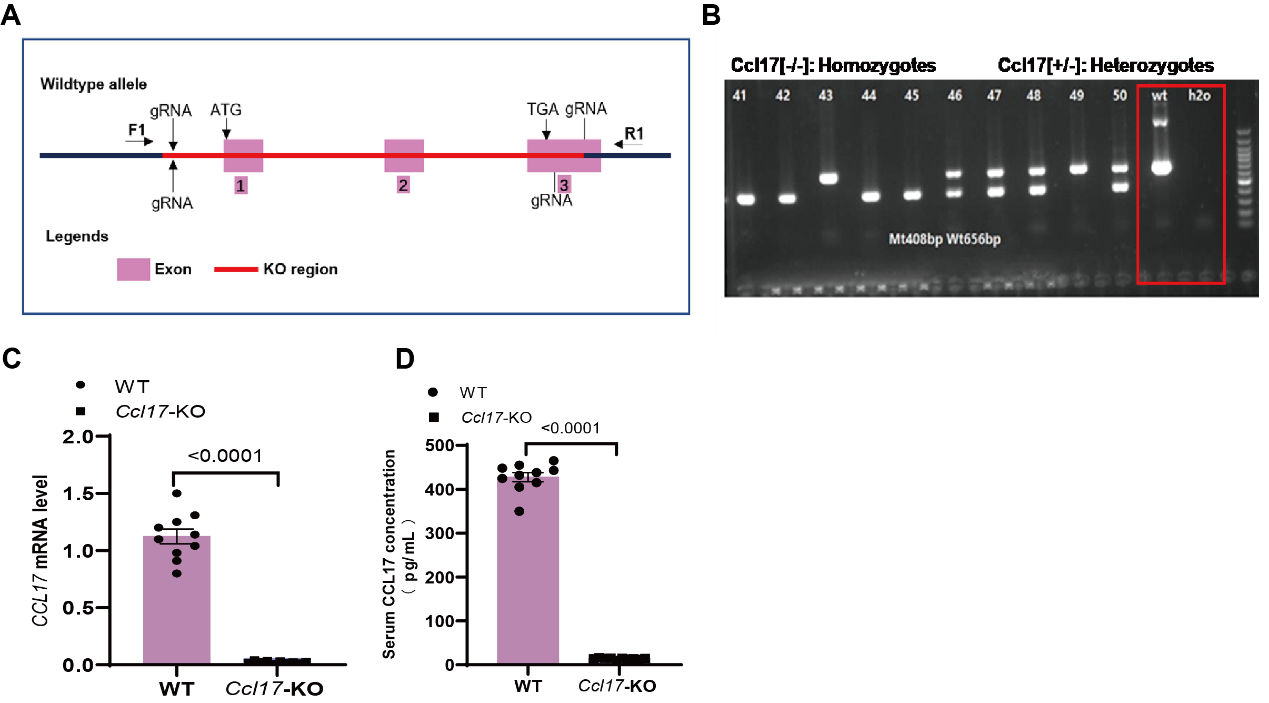

**Supplementary Figure 2. Generation and validation of global Ccl17-knockout mice.**
(A) Schematic of the CRISPR/Cas9-mediated *Ccl17* deletion strategy. The targeted region spanning exons 1–3 is indicated in red. (B) Representative PCR genotyping of WT, heterozygous, and homozygous *Ccl17*-KO mice. PCR products of 656 and 408 base pairs correspond to the WT and targeted alleles, respectively. Water was used as the negative control. (C) Relative cardiac *Ccl17* mRNA expression in WT and *Ccl17*-KO mice. (D) Serum CCL17 concentrations measured by enzyme-linked immunosorbent assay in WT and Ccl17-KO mice. n=8 mice per genotype in (C,D). Data are presented as mean ± SEM. Statistical significance was assessed using a two-tailed unpaired Student’s t test; P<0.0001. Each symbol represents one mouse.

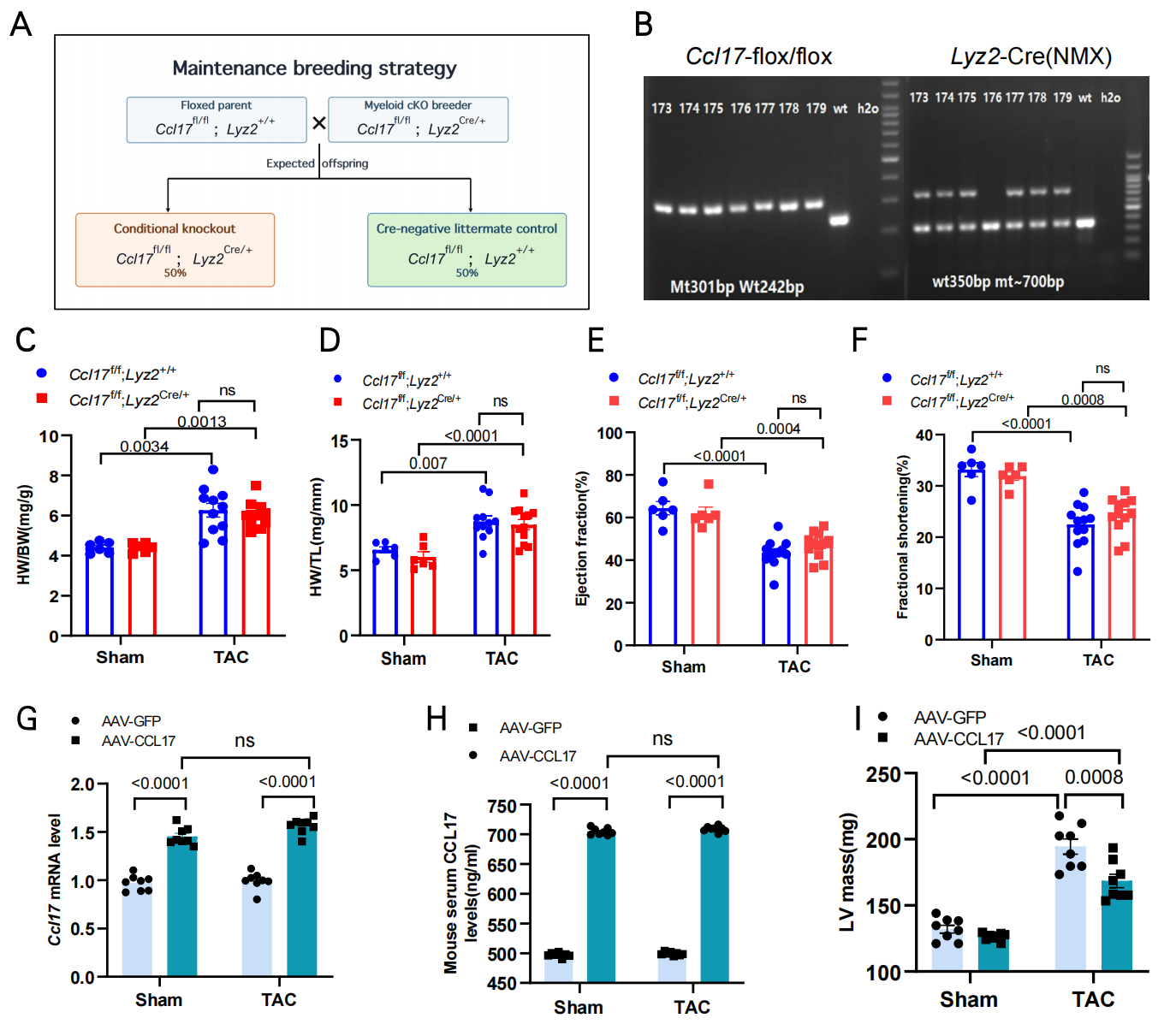

**Supplementary Figure 3. Generation and phenotypic assessment of myeloid-restricted Ccl17 knockout mice and validation of AAV-mediated CCL17 augmentation.**
(A) Schematic of the maintenance breeding strategy used to generate myeloid-restricted Ccl17 conditional knockout mice (Ccl17^fl/fl^;Lyz2^Cre/+^) and Cre-negative littermate controls (Ccl17^fl/fl^;Lyz2^+/+^). (B) Representative agarose gel electrophoresis showing PCR genotyping of the floxed Ccl17 allele and the Lyz2-Cre allele. Expected amplicon sizes are indicated beneath the gels; wt, wild-type control; H₂O, no-template control. (C–F) Heart weight-to-body weight ratio (HW/BW; C), heart weight-to-tibia length ratio (HW/TL; D), left ventricular ejection fraction (E) and fractional shortening (F) in Ccl17^fl/fl^;Lyz2^+/+^ and Ccl17^fl/fl^;Lyz2^Cre/+^ mice 6 weeks after sham or transverse aortic constriction (TAC) surgery (n = 8 mice per Sham group and n = 12 mice per TAC group). (G) Relative cardiac Ccl17 mRNA expression in mice treated with AAV-F4/80-GFP or AAV-F4/80-CCL17 at the study endpoint, 6 weeks after sham or TAC surgery (n = 8 mice per group). (H) Serum CCL17 concentrations measured by enzyme-linked immunosorbent assay in the indicated AAV-treated groups at the study endpoint (n = 6 mice per group). (I) Left ventricular (LV) mass assessed by echocardiography 6 weeks after sham or TAC surgery in mice treated with AAV-F4/80-GFP or AAV-F4/80-CCL17 (n = 8 mice per group). Data are presented as mean ± SEM. Each symbol represents one mouse. Statistical significance was assessed using two-way ANOVA followed by Tukey’s multiple-comparisons test. Exact P values are indicated in the panels; ns, not significant.

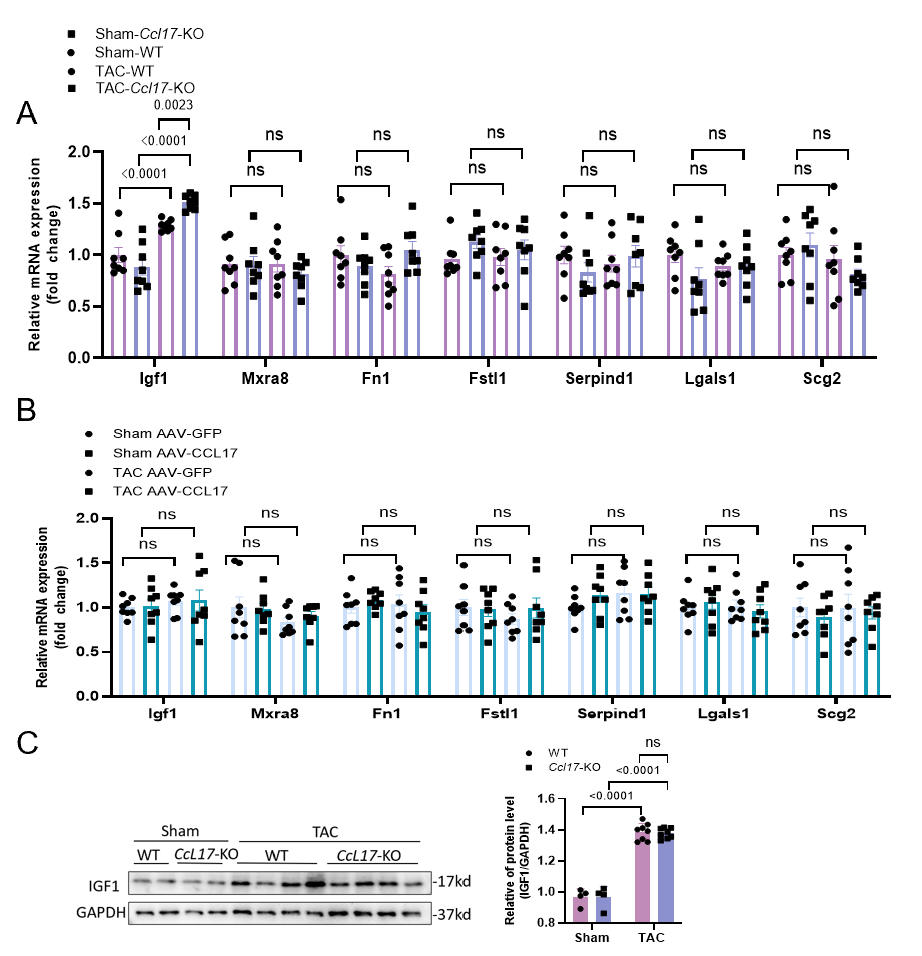

**Supplementary Figure 4. Experimental validation of candidate genes from the IGFBP-associated pathway identified by cardiac RNA sequencing.** (A) Relative cardiac mRNA expression of *Igf1*, *Mxra8*, *Fn1*, *Fstl1*, *Serpind1*, *Lgals1*, and *Scg2* in wild-type (WT) and *Ccl17*-knockout (*Ccl17*-KO) mice 6 weeks after sham or transverse aortic constriction (TAC) surgery (n = 8 mice per group). (B) Relative cardiac mRNA expression of the same candidate genes in mice treated with AAV-F4/80-GFP or AAV-F4/80-CCL17 6 weeks after sham or TAC surgery (n = 8 mice per group). (C) Representative immunoblots and densitometric quantification of cardiac IGF1 protein abundance in WT and *Ccl17*-KO mice 6 weeks after sham or TAC surgery. GAPDH served as the loading control (n = 4 mice per sham group and n = 8 mice per TAC group). Data are presented as mean ± SEM. Each symbol represents one mouse. For each gene in (A) and (B) and for IGF1 protein abundance in (C), statistical significance was assessed using two-way ANOVA followed by Tukey’s multiple-comparisons test. Exact *P* values are indicated in the panels; ns, not significant.

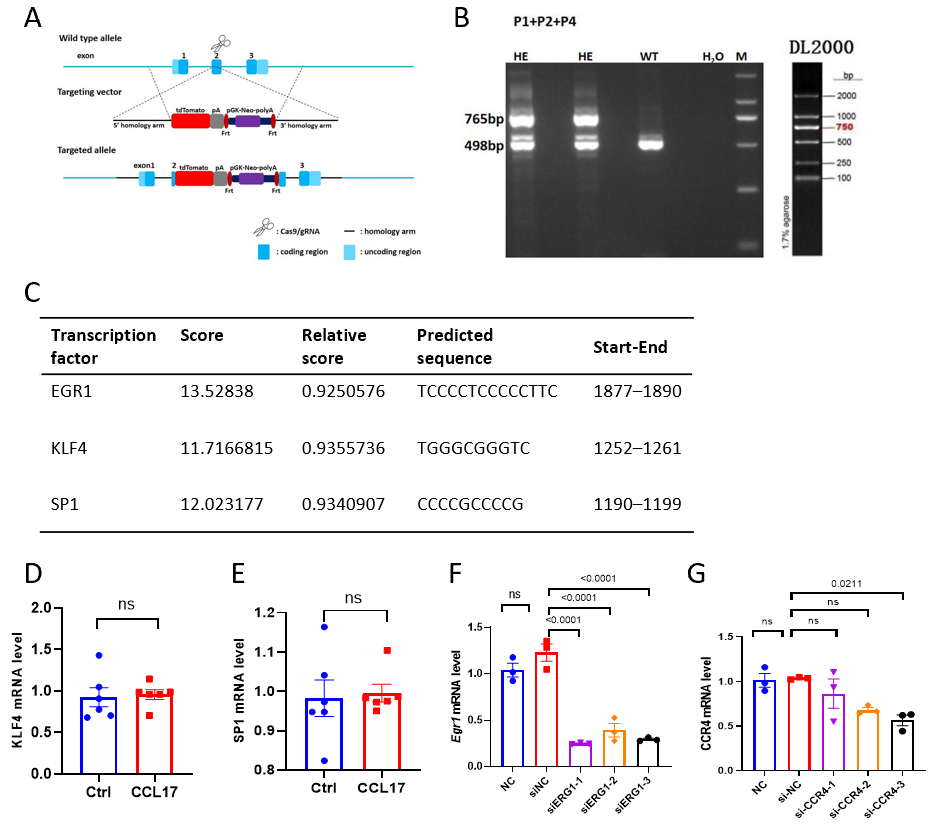

**Supplementary Figure 5. Validation of the *Ccl17*-tdTomato reporter mouse and molecular tools used to investigate CCR4–EGR1 signalling.
(A)** Schematic of the strategy used to generate *Ccl17*-tdTomato reporter mice. The targeted allele contains tdTomato–poly(A) and FRT-flanked PGK-Neo–poly(A) cassettes inserted into the *Ccl17* locus. **(B)** Representative agarose gel electrophoresis of PCR products generated using primers P1, P2 and P4 for genotyping *Ccl17*-tdTomato mice. The targeted and wild-type alleles produced bands of 765 and 498 bp, respectively. HE, heterozygous; WT, wild type; H₂O, no-template control; M and DL2000, DNA molecular weight markers. **(C)** Candidate transcription factors predicted to bind the rat *Igfbp6* promoter using the JASPAR database, showing the raw score, relative score, predicted binding sequence and position within the analysed promoter sequence. **(D,E)** Relative *Klf4* **(D)** and *Sp1* **(E)** mRNA expression in neonatal rat cardiac fibroblasts (NRCFs) treated with vehicle or recombinant CCL17 (rCCL17; 100 ng/mL; n=6 independent experiments per group). **(F)** Relative *Egr1* mRNA expression in rCCL17-treated NRCFs following no transfection (NC) or transfection with negative-control siRNA (siNC) or one of three EGR1-targeting siRNAs (siEGR1-1–siEGR1-3; n=3 independent experiments per group). **(G)** Relative *Ccr4* mRNA expression in rCCL17-treated NRCFs following no transfection (NC) or transfection with siNC or one of three CCR4-targeting siRNAs (siCCR4-1–siCCR4-3; n=3 independent experiments per group). Individual symbols represent independent experiments. Data are presented as mean±SEM. Comparisons between two groups in **(D,E)** were performed using two-tailed unpaired Student’s *t* tests. Comparisons among multiple groups in (F,G) were performed using one-way ANOVA followed by Tukey’s multiple-comparisons test. Exact *P* values are indicated in the panels; ns, not significant.

| **Abbreviation** | **Full term** |
| --- | --- |
| **AS** | aortic stenosis |
| **CCL17** | C-C motif chemokine ligand 17 |
| **CCR4** | C-C chemokine receptor 4 |
| **EF** | ejection fraction |
| **EGR1** | early growth response 1 |
| **IGF** | insulin-like growth factor |
| **IGF1** | insulin-like growth factor 1 |
| **IGFBP** | insulin-like growth factor-binding protein |
| **IGFBP6** | insulin-like growth factor-binding protein 6 |
| **LV** | left ventricular |
| **TAC** | transverse aortic constriction |
| **TARC** | thymus and activation-regulated chemokine |
| **TGF-β1** | transforming growth factor-β1 |
| **α-SMA** | α-smooth muscle actin |
| **NPX** | normalized protein expression |

| Gene | Forward sequence（5′–3′） | Reverse sequence（5′–3′） |
| --- | --- | --- |
| Human CCL17 | ATTCAAAACCAGGGTGTCTCC | TGGATGTGCTGCAGAGAAG |
| Human GAPDH | TTGTCAAGCTCATTTCCTGGT | TCTCTCTTCCTCTTGTGCTCT |
| Mouse Ccl17 | TGCTGCCTGGATTACTTCAAA | CTGGACAGTCAGAAACACGAT |
| Mouse Gapdh | TTGTCAAGCTCATTTCCTGGT | GATAGGGCCTCTCTTGCTCA |
| Mouse Nppa | CTTCCTCGTCTTGGCCTTTT | GTGGTCTAGCAGGTTCTTGAA |
| Mouse Nppb | GTCCAGCAGAGACCTCAAAAT | CAACAACTTCAGTGCGTTACA |
| Mouse Myh7 | CATCCTCATCACCGGAGAATC | GGCGGCAATAACAGCAAAATA |
| Mouse Postn | TCCTGATTCTGCCAAACAAGT | GTGTACTCTCCATCTGGCTTC |
| Mouse Col1a1 | AGACATGTTCAGCTTTGTGGA | ACTTCAGGGATGTCTTCTTGG |
| Mouse Col3a1 | TGGCTGTACTAAACACACTGG | GGGTGCGATATCTATGATGGG |
| Mouse Igfbp6 | GAAAGCAGCAGTGTCGTTC | CTTCCTTGACCATCTGGAGAC |
| Mouse Igf1 | TTACTTCAACAAGCCCACAGG | GCACAGTACATCTCCAGTCTC |
| Mouse Mxra8 | CTGCTCATCACTGTAGTCCTG | CACATCCTTCCCCTTTGACTT |
| Mouse Fn1 | AGCACTACCCCAGATATCACT | AACACTGACGTTGTACTCCAG |
| Mouse Fstl1 | CAAGACCTACCTCAACCACTG | CGCAGACTTCTTTTCTTTGCA |
| Mouse Serpind1 | AGCATGACAAACAGAACTCGA | CATGGACTTAAGGACCTCCAC |
| Mouse Lgals1 | AATCATGGCCTGTGGTCTG | CAGGTTGTTGCTGTCTTTTCC |
| Mouse Scg2 | AGAAAACGGAGAAGAAAGCCA | CCTCGAGTATTATCCGCATCC |
| Rat Igfbp6 | CGCAGACACTTGGATTCAGTA | GGAACGACACTGCTGCTT |
| Rat Ccr4 | CTTCCGTGACGCTTTGTAATG | GGACTCCCTGACTGAAACAAA |
| Rat Egr1 | CGGCAATAACAGCAGCAG | CAGAAAAGGACTCTGTGGTCA |
| Rat Gapdh | GGTTGTCTCCTGCGACTTCA | TGGTCCAGGGTTTCTTACTCC |
| Rat Klf4 | CACACAGGCGAGAAACCTTA | GTAGTGCCTGGTCAGTTCATC |
| Rat Sp1 | CATGAGCGACCAAGATCACTC | TGCTTCGAGTCTGAGAAAAGG |

**Supplementary Table 1. Primer sequences used for RT–qPCR and genotyping**

| **Antibody** | **Catalog** | **Manufacturer** |
| --- | --- | --- |
| Anti-TARC/CCL17 antibody | Ab182793 | Abcam |
| Alpha-Smooth Muscle Actin Monoclonal Antibody | 14-9760-82 | Invitrogen |
| GAPDH Antibody Rabbit Polyclonal | ES-Pab001/100 | YISHAN Biotechnology |
| IGFBP6 antibody | DF8959 | Affinity |
| IGF1 antibody | DF6096 | Affinity |
| CCR4 Antibody | DF101206 | Affinity |
| Alpha-Smooth Muscle Actin Rabbit mAb | R380653 | Zenbio |
| Anti-RFP mAb Cocktail | M208-3 | MBL |
| Anti-beta Tubulin Antibody | Ab6046 | Abcam |

**Supplementary Table 2. Antibodies used in this study**

| **Sequence name** | **Species** | **Sense sequence** | | **Antisense sequence** | | **Sense length (nt)** | | **Antisense length (nt)** |
| --- | --- | --- | --- | --- | --- | --- | --- | --- |
| **siRNA-mrGAPDH** | Mouse /Rat | UCACUCAAGAUUGUCAGCAAUGCAU | AUGCAUUGCUGACAAUCUUGAGUGA | | 25 | | 25 | |
| **siRNA-NC** | Not specified | UUCUCCGAACGUGUCACGUTT | ACGUGACACGUUCGGAGAATT | | 21 | | 21 | |
| **rat-Ccr4(ID:171054)-sirna-138** | Rat | GGAUGAAGCCGCGUACAAUTT | AUUGUACGCGGCUUCAUCCTT | | 21 | | 21 | |
| **rat-Egr1(ID:24330)-sirna-1920** | Rat | GGAGCGCGAGAGACAAUAATT | UUAUUGUCUCUCGCGCUCCTT | | 21 | | 21 | |
| **rat-Klf4(ID:114505)-sirna-1308** | Rat | CGAAGAGUUCUCAUCUCAATT | UUGAGAUGAGAACUCUUCGTT | | 21 | | 21 | |
| **rat-Sp1(ID:24790)-sirna-7042** | Rat | GGUUGUGUGCUUUCCUAUATT | UAUAGGAAAGCACACAACCTT | | 21 | | 21 | |

**Supplementary Table 3. Sequences of siRNAs used in this study**

| **Site** | **JASPAR matrix ID** | **Position relative to TSS (bp)** | **Strand** | **WT sequence（5′–3′）** | **Mutant sequence（5′–3′）** |
| --- | --- | --- | --- | --- | --- |
| **S1** | **MA0162.5** | **−1125 to −1116** | **+** | **CCGCCCACCC** | **AAAAAAAAAA** |
| **S2** | **MA0162.5** | **−799 to −790** | **+** | **CCGCCCACCC** | **AAAAAAAAAA** |
| **S3** | **MA0162.2** | **−124 to −111** | **+** | **TCCCCTCCCCCTTC** | **AAAAAAAAAAAAAA** |

A. **Predicted EGR1-binding sites and mutations**

| **Construct** | **Promoter region relative to TSS (bp)** | | **Insert length (bp)** | **Mutated site(s)** | **Vector** |
| --- | --- | --- | --- | --- | --- |
| **WT** | | **−2000至+2** | **2002** | **None** | **pGL3-Basic** |
| **Mut-S1** | | **−2000至+2** | **2002** | **S1** | **pGL3-Basic** |
| **Mut-S2** | | **−2000至+2** | **2002** | **S2** | **pGL3-Basic** |
| **Mut-S3** | | **−2000至+2** | **2002** | **S3** | **pGL3-Basic** |
| **Mut**  **(S1+S2+S3)** | | **−2000至+2** | **2002** | **S1+S2+S3** | **pGL3-Basic** |

B. **Igfbp6 promoter reporter constructs**

**Supplementary Table 4. Rat Igfbp6 promoter sequences and EGR1-binding-site mutations used in dual-luciferase reporter assays**
